# Characterizing patterns of diffusion tensor imaging variance in aging brains

**DOI:** 10.1101/2023.08.22.23294381

**Authors:** Chenyu Gao, Qi Yang, Michael E. Kim, Nazirah Mohd Khairi, Leon Y. Cai, Nancy R. Newlin, Praitayini Kanakaraj, Lucas W. Remedios, Aravind R. Krishnan, Xin Yu, Tianyuan Yao, Panpan Zhang, Kurt G. Schilling, Daniel Moyer, Derek B. Archer, Susan M. Resnick, Bennett A. Landman, the Alzheimer’s Disease Neuroimaging Initiative, the BIOCARD Study team

## Abstract

**Purpose:** As large analyses merge data across sites, a deeper understanding of variance in statistical assessment across the sources of data becomes critical for valid analyses. Diffusion tensor imaging (DTI) exhibits spatially varying and correlated noise, so care must be taken with distributional assumptions. Here we characterize the role of physiology, subject compliance, and the interaction of subject with the scanner in the understanding of DTI variability, as modeled in spatial variance of derived metrics in homogeneous regions.

**Approach:** We analyze DTI data from 1035 subjects in the Baltimore Longitudinal Study of Aging (BLSA), with ages ranging from 22.4 to 103 years old. For each subject, up to 12 longitudinal sessions were conducted. We assess variance of DTI scalars within regions of interest (ROIs) defined by four segmentation methods and investigate the relationships between the variance and covariates, including baseline age, time from the baseline (referred to as “interval”), motion, sex, and whether it is the first scan or the second scan in the session.

**Results:** Covariate effects are heterogeneous and bilaterally symmetric across ROIs. Inter-session interval is positively related (*p* ≪ 0.001) to FA variance in the cuneus and occipital gyrus, but negatively (*p* ≪ 0.001) in the caudate nucleus. Males show significantly (*p* ≪ 0.001) higher FA variance in the right putamen, thalamus, body of the corpus callosum, and cingulate gyrus. In 62 out of 176 ROIs defined by the Eve type-1 atlas, an increase in motion is associated (*p* < 0.05) with a decrease in FA variance. Head motion increases during the rescan of DTI (Δ*μ* = 0.045 millimeters per volume).

**Conclusions:** The effects of each covariate on DTI variance, and their relationships across ROIs are complex. Ultimately, we encourage researchers to include estimates of variance when sharing data and consider models of heteroscedasticity in analysis. This work provides a foundation for study planning to account for regional variations in metric variance.

## 1 Introduction

Large datasets enable exploration of questions that would be impractical with smaller- or moderate-sized datasets while giving rise to the development and application of deep learning models which can assimilate complex data. One prevalent challenge is that large datasets often comprise samples aggregated from distinct sources at different time points using diverse technologies, causing data heterogeneity, experimental variations, and statistical biases if the analysis is not executed appropriately.^1^ In such scenarios, understanding the variance and variability of data is of great importance. The general linear model^2^, a structured and widely used framework for relationship modeling, allows us to illustrate the importance. The general linear model is assessed through regression. A linear regression can be expressed by Y = Xβ + ε, where the response variable Y, the covariate matrix X, and the regression coefficients β are conventionally represented in matrix forms given by:

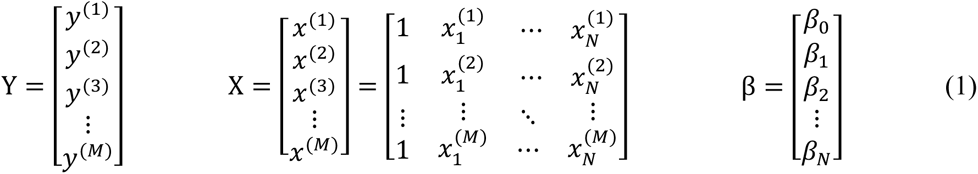

where we use *M* to denote the number of samples, and *N* to denote the number of independent variables. The error term ε is given by:

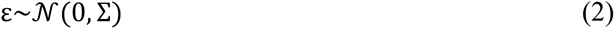

where Σ represents the covariance matrix. If we assume the errors are uncorrelated, Σ is simplified to a diagonal matrix. We can simplify estimation of Eq. (1) with a whitening matrix^3^, W:

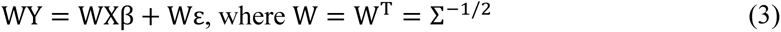

Note Wε∼𝒩(0, I*_M_*), where I*_M_* denotes the identity matrix of dimension *M* × *M*. We illustrate the practical importance of understanding the variance structure for reducing statistical errors (Fig. 1).

**Fig. 1.**
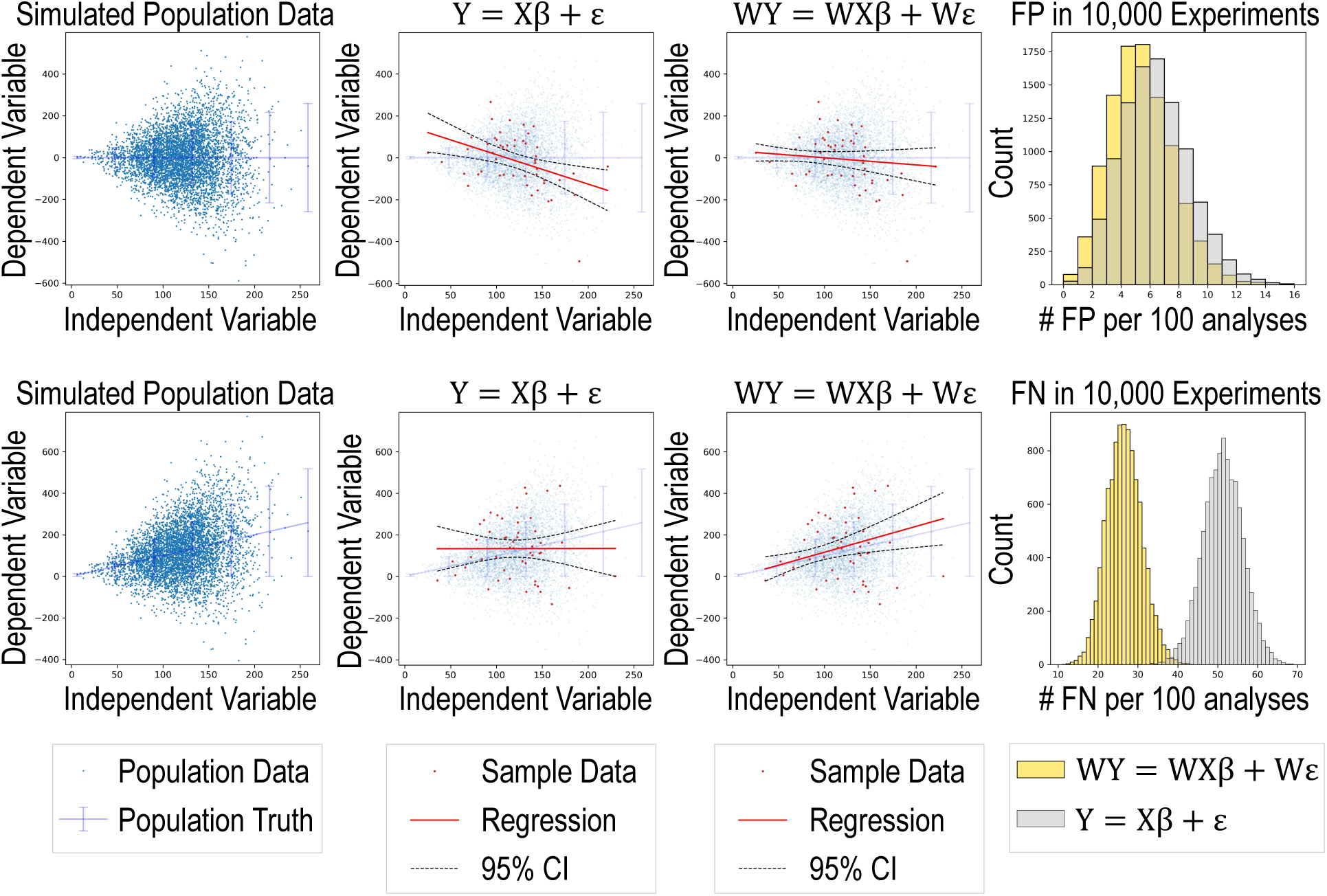
Simulation shows that applying the whitening matrix to the standard linear regression equation reduces the number of false positives (FP) and false negatives (FN) under heteroskedasticity. In the top row, the population truth has zero slope. In data sampled from the synthetic population data, ordinary least square (OLS) regression using the standard equation generates FP, while the solution with whitening, W, does not falsely reject the null hypothesis (the horizontal line). After 10,000 experiments, the FP frequency is lower with whitening, centering at 5 per 100. In the second row, the population truth has a positive slope. In data sampled from the synthetic population data, OLS regression using the standard equation generates FN, while the solution with whitening, W, does not. After 10,000 experiments, the FN frequency with whitening is half that of the one without whitening.

Diffusion tensor imaging (DTI)^4–6^ is a modeling approach used in diffusion-weighted imaging ^7–9^, a variant of conventional magnetic resonance imaging (MRI) based on the tissue water diffusion rate.^10^ DTI allows for visualization and measurement of the degree of anisotropy and structural orientation of fibers in the brain and has been widely used in studies.^11–13^ DTI is inherently subject to low signal-to-noise ratio, and the noise structure exhibits spatial variability and correlation, primarily attributed to fast imaging and noise suppression techniques.^14,15^ Understanding the statistical nature of DTI variance or noise has been proven to be beneficial for diffusion tensor estimation^15,16^, outlier detection^17^, reproducibility assessment^18–20^.

Methods have been proposed for DTI variance (or noise) estimation, among which we recognize four types. The first type requires multiple repeated acquisitions (therefore, we refer to it as the “multiple acquisition method”). After the repeated acquisitions, we could take the standard deviation of the measurements, or perform data resampling, such as bootstrap or jackknife,^21^ to quantify uncertainties of DTI parameters. The second type involves two repeated acquisitions, which we call the “scan-rescan method”. We compute the difference between the images from each acquisition and then calculate the standard deviation of the difference across the space.^22,23^ Note that the standard deviation must be divided by the square root of 2 to account for the combination of two random variables (noise in each image). The third and fourth types are used when we have only one acquisition. For the third type, we select a homogeneous ROI and compute the standard deviation of the measurements within this ROI (“ROI-based method”).^24^ The fourth type, often referred to as “model-based resampling”, involves fitting a model (e.g., diffusion tensor) locally to the observed data. The residuals from the fitted model, along with the original observed data, are then used by data resampling techniques such as wild bootstrap to generate random subsets.^21,25^ From each subset, we obtain an estimate of a specific parameter. Across all subsets, we get the distribution of the estimates and thus quantify the uncertainty of the DTI parameter. The first type (multiple acquisition method) makes no assumptions about the noise properties at the cost of requiring multiple acquisitions (for each diffusion gradient in the DTI scenario).^21^ The second type (scan-rescan method) assumes that the noise is constant across space, or across the region from which the standard deviation is computed.^23^ The third method (ROI-based method) assumes that both the noise and the signal are constant across the ROI.^24^ The fourth method (model-based resampling) assumes that the non-constant variance of measured signals can be captured by the chosen model.^21^ In this study, we choose the ROI-based method for estimating noise across brain regions in DTI. This is because it does not require repeated acquisitions, and one advantage is that we can compute noise from each individual scan within an imaging session, enabling inter-scan comparisons. Also, this method can be applied to datasets that do not have scan-rescan data available, thus enabling validation of our findings using other datasets. Secondly, by using an individual scan for noise estimation, we can mitigate errors caused by motion and inter-scan misalignment of the brain, which could be problematic when using the multiple acquisition method or the scan-rescan method. Thirdly, we want to avoid using the assumption of the fourth type (model-based resampling).^21^

Up to this point, we have been using the terms “variance” and “noise” interchangeably. In the following text, we use “variance” when referring to the measure of the dispersion of data, and “noise” when referring to the imaging noise such as MRI noise—primarily caused by thermal fluctuations and electrical noise—to avoid misinterpretation.

To gain a better understanding of DTI variance or noise, it is important to characterize the role of physiology, subject compliance, and the interaction between the subject and the scanner. Our approach is driven by two fundamental questions (Fig. 2): Which factors are associated with DTI variance? Where and how does this association manifest? We assess variance of DTI scalars, including fractional anisotropy (FA), axial diffusivity (AD), mean diffusivity (MD), and radial diffusivity (RD), within ROIs, and investigate the associations between the variance and covariates, including baseline age, time from the baseline (referred to as “interval”), motion, sex, and whether it is the first or the second scan within the session, using linear mixed effects models^26^.

**Fig. 2.**
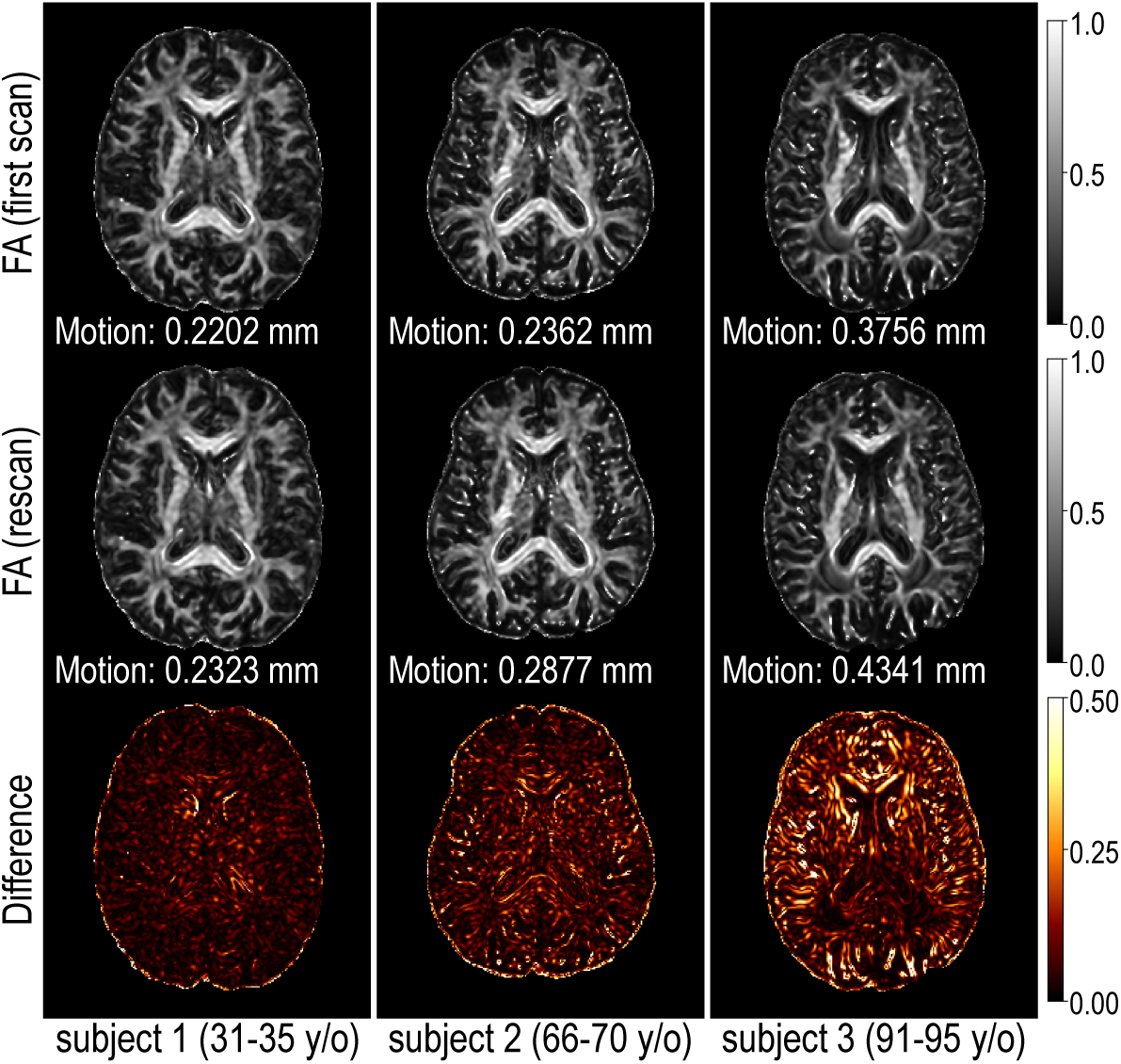
We observe that the noise (approximated by the difference between the scan and rescan acquired within the same imaging session) in DTI scalar images, such as fractional anisotropy (FA) images, generally increase with age. (Subjects’ ages are grouped into five-year bins to respect privacy.) But motion is also considered to increase with age.67,68 We would like to know: Which factor is associated with DTI variance? Where and how does this association manifest?

## 2 Methods

We use the PreQual^27^ pipeline for preprocessing and quality assurance of the DTI data. PreQual is an end-to-end pipeline that applies denoising, inter-scan normalization, susceptibility-induced distortion correction, eddy current-induced distortion correction, inter-volume motion correction, slice-wise signal dropout imputation, and more. PreQual also provides a summary of the data and preprocessing in a PDF report for more efficient quality assurance.

We use data acquired from the neuroimaging substudy of the Baltimore Longitudinal Study of Aging (BLSA).^28,29^ The BLSA is an extensive, ongoing research project that began in 1958, enrolling healthy volunteers aged 20 years and older to study normal aging through a longitudinal approach by following participants for their entire lives. We consider all subjects with at least one session comprising both T1-weighted (T1w) magnetization-prepared rapid gradient-recalled echo (MPRAGE) MRI data and DTI data. We exclude 49 DTI images exhibiting one or more of the following characteristics according to their potential impact on subsequent analyses:

1. The presence of extreme susceptibility-induced distortion, motion artifacts, or eddy currents that resists correction.
2. The failure of the preprocessed data to be fitted by the tensor model.
3. An exceptionally low signal-to-noise ratio in the FA and MD images.

The exclusion of these cases results in the dataset depicted in Fig. 3. We identify 1035 subjects (562 F/ 473 M, 22.4 to 94.4 y/o at baseline) with 2751 sessions (1497 F/ 1254 M). Detailed demographic information can be found in the supplementary material. The 2751 sessions of MRI data were acquired by four different scanners, including a 1.5 Tesla Philips Intera scanner (scanner A, 83 sessions) at the Kennedy Krieger Institute (KKI), two 3 Tesla Philips Achieva scanners using the same platform and protocol (scanner B and C, 16 sessions and 59 sessions, respectively) at the KKI, and a 3 Tesla Philips Achieva scanner (scanner D, 2593 sessions) at the National Institute on Aging. Detailed scanner information and protocol is provided in Table 1, which was previously reported^30^. Among the 1035 subjects, 59 switched to a different scanner in follow-up scans. During subsequent sessions, 4 female subjects and 10 male subjects were diagnosed with Alzheimer’s disease, while the remaining subjects remained cognitively normal throughout all sessions.

**Fig. 3.**
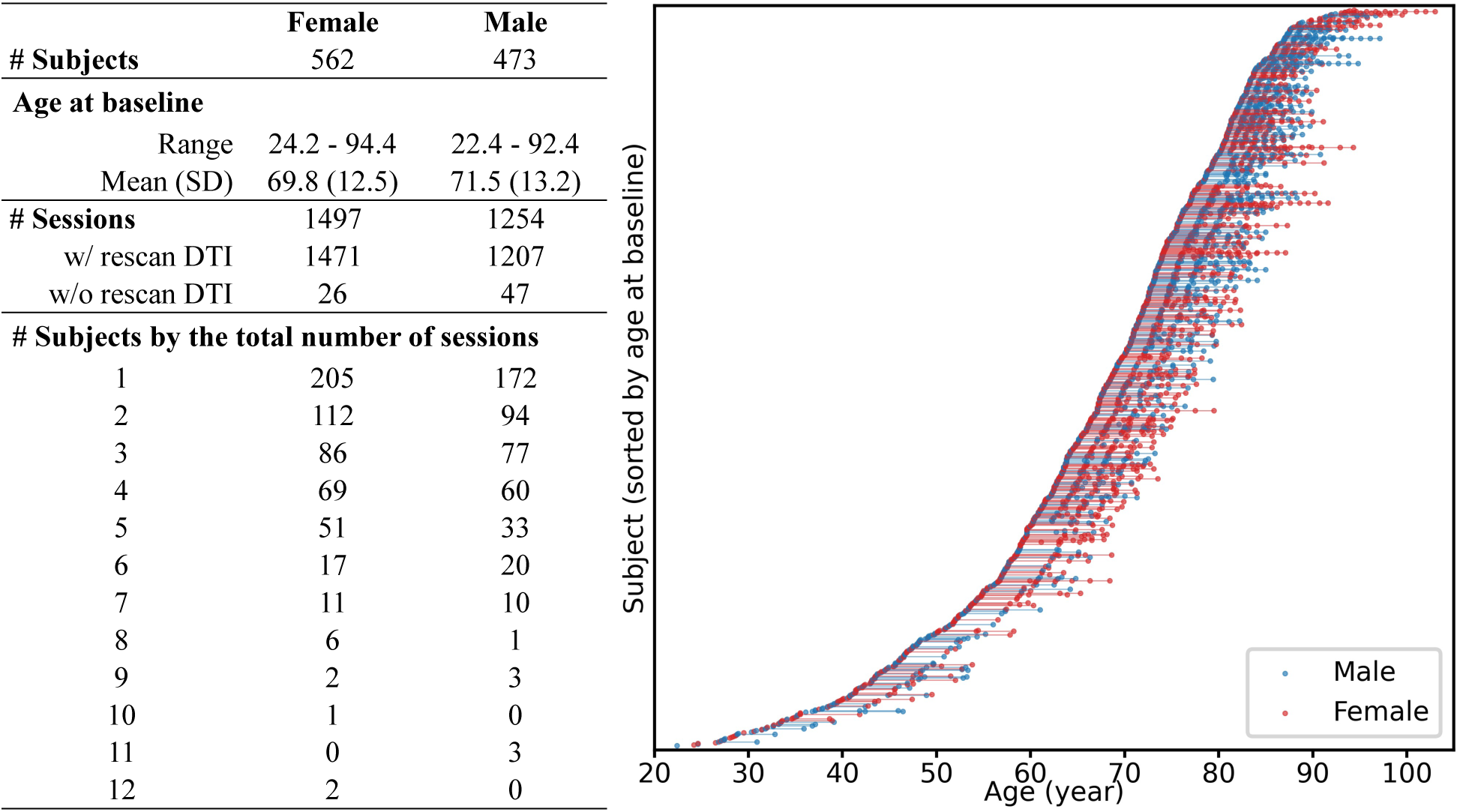
The BLSA dataset we use has a slight imbalance between the number of females and males, but it is well-matched and appropriate for our research objectives in other aspects: i) the age ranges of females and males align closely; ii) rescan DTI data were acquired in most sessions, enabling inter-scan comparisons; iii) the distributions of sessions of females and males align closely.

**Table 1.**
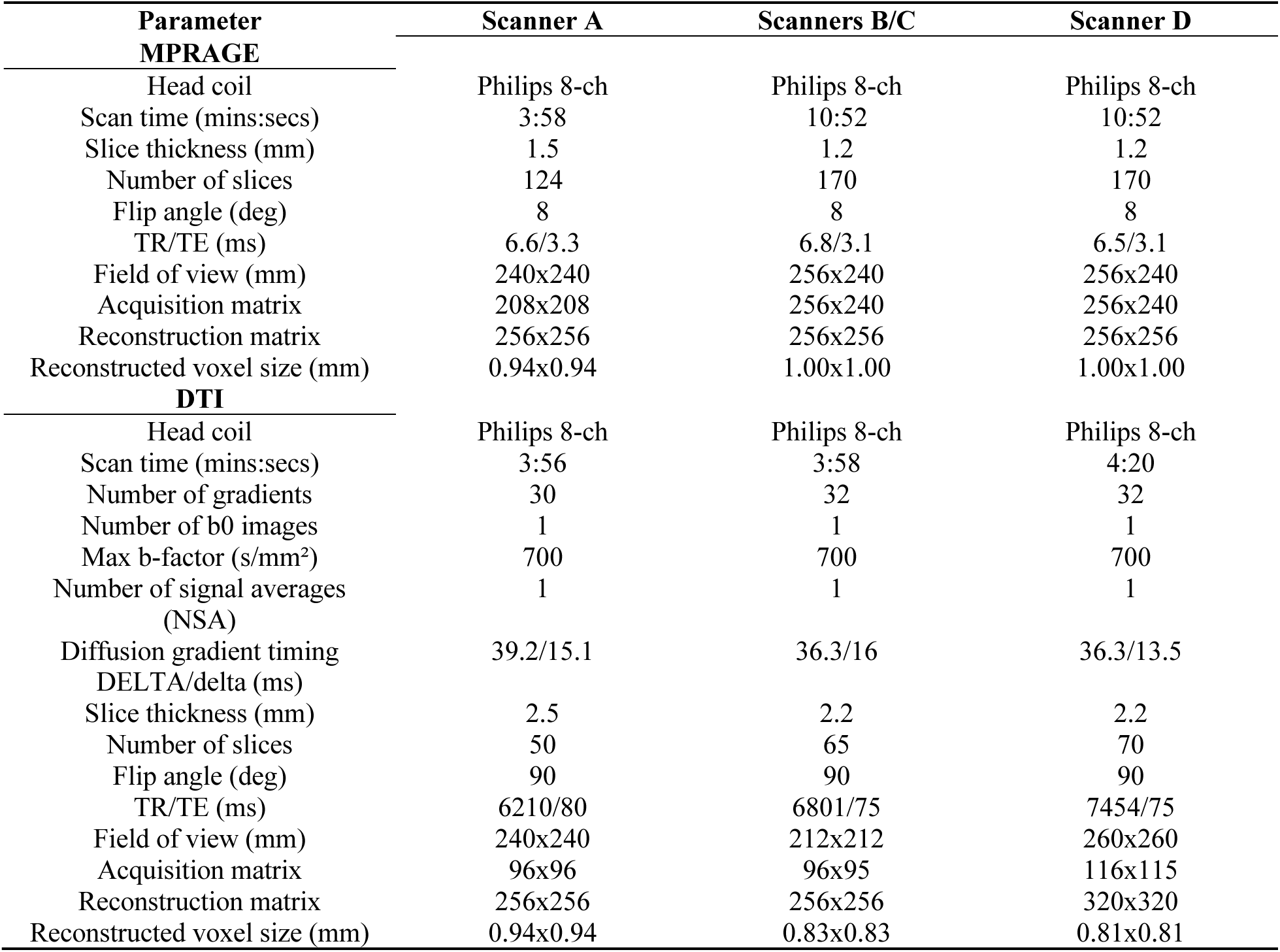
Protocol for the T1w MPRAGE scan and DTI scan.

### 2.1 ROI-Based DTI Variance Estimation

We use a registration-based approach for brain segmentation in the b0 (minimally weighted) volume (Fig. 4). We initiate the process with brain segmentations for the T1w images obtained through manual parcellations provided by the JHU-MNI-ss atlas (“Eve atlas”)^31,32^ and automated whole-brain segmentation by SLANT^33^. For the Eve atlas, there are three types of parcellations available, each with different regional focus.^31^ For SLANT segmentation, labels for 132 regions covering the whole brain are provided.^33^ We use the method by Hansen et al.^34^ to transfer these labels from T1w to b0 space. After label transferring, we manually review the segmentation to see if the labels align with the anatomical regions.

**Fig. 4.**
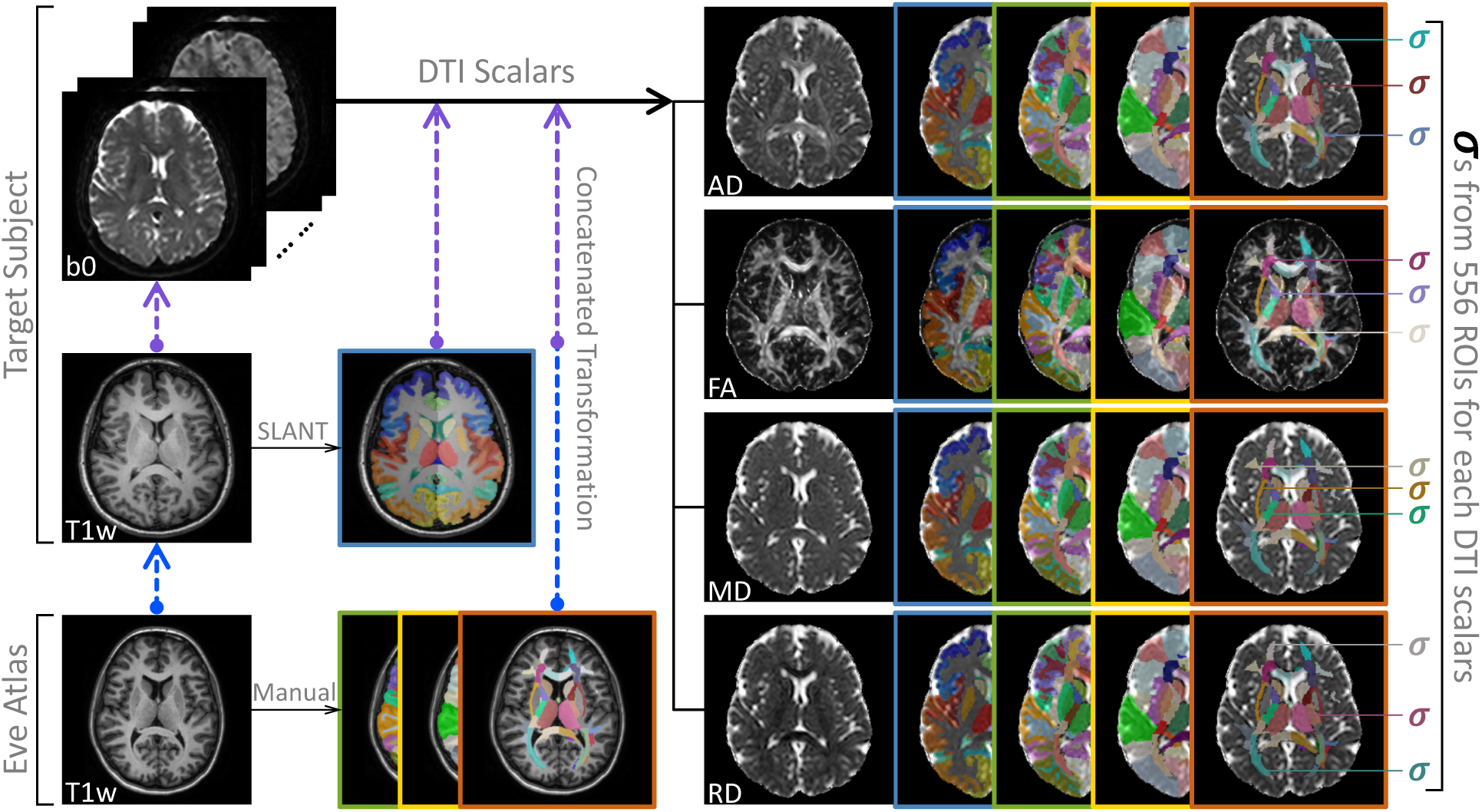
Brain segmentation labels are obtained using the SLANT segmentation of the target subject’s T1w image, and using three types of manual parcellations provided by the Eve atlas. To generate transformation matrices for transferring these labels to DTI scalar images, intra- and inter- modality registrations are performed. Standard deviations of DTI scalars within each ROI are computed.

### 2.2 Linear Mixed-Effects Model

We use linear mixed-effects models^26^ to analyze the association between DTI scalar standard deviation and covariates. (R program, version 4.2.2 ^35^; Ubuntu 20.04.5 LTS; R package lme4, version 1.1.31 ^36^; R package lmerTest, version 3.1.3 ^37^.)

We study linear mixed-effects models of the form:

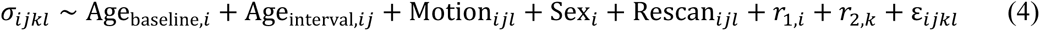

where *σ_ijkl_* represents the standard deviation of a DTI scalar (FA, AD, MD, or RD) in a specific brain region of subject *i* at session *j* via scanner *k* in acquisition *l*, Age_baseline,*i*_ (hereafter referred to as “baseline”) is the age of subject *i* at baseline session (unit: decade), Age_interval,*i*,*j*-_ (hereafter referred to as “interval”) is the time between the current session, *j*, and the baseline session (unit: decade). Motion_,-/_ is a scalar value reflecting the degree of head movement of subject *i* at session *j* during acquisition *l* (calculated based on eddy movement, unit: millimeters)^38^, Sex, is the gender of subject *i* (0 for female and 1 for male), and Rescan_,-/_ is a binary variable indicating if the acquisition *l* is the first scan (coded 0) or the rescan (coded 1) of session *j*. We consider subject and scanner as two random intercepts, respectively denoted by *r*_1,*i*_ and *r*_2*k*,._. Prior to fitting the models, we standardize the dependent variable *σ*. The results for the models are based on the standardized *σ*.

We have a total of 2224 models, derived from the four DTI scalars (FA, AD, MD, or RD), across varying ROIs defined by Eve Type 1 (176 ROIs), Eve Type 2 (130 ROIs), Eve Type 3 (118 ROIs),^31,32^ and SLANT (132 ROIs)^33^. Each model starts with a full model, with all fixed effects and random effects, followed by an implementation of backward model selection.^37^ The p-values for the fixed-effect terms are calculated based on the associated F tests.^37^ To account for multiple comparisons, we adjust the p-values across the pairs of DTI scalar and ROI for a false discovery rate (FDR) of 0.05 using the Benjamini-Hochberg method.^39^

## 3 Results

The magnitude and direction of the effects of each covariate on DTI variance exhibit heterogeneous patterns across ROIs (Fig. 5). Specifically, interval is positively related to FA variance in ROIs such as the cuneus, middle occipital gyrus, superior occipital gyrus, medulla, precuneus white matter, but negatively related in ROIs such as the caudate nucleus, posterior thalamic radiation, and superior fronto-occipital fasciculus (Table 2). Males have higher FA variance in the right putamen, thalamus, body of corpus callosum, and cingulum (cingulate gyrus), but lower FA variance in the middle frontal gyrus (Table 3). In the right inferior temporal gyrus, an increase of 1 millimeter in motion is associated with an increase of 2.211 standard deviations in the z-scored standard deviation (*σ*) of FA values (*β* = 2.211, *p* ≪ 0.001). Interestingly and counterintuitively, in several ROIs, including the medulla, middle occipital white matter, cingulum (cingulate gyrus), an increase in motion is linked with a decrease in FA variance (Table 2).

**Fig. 5.**
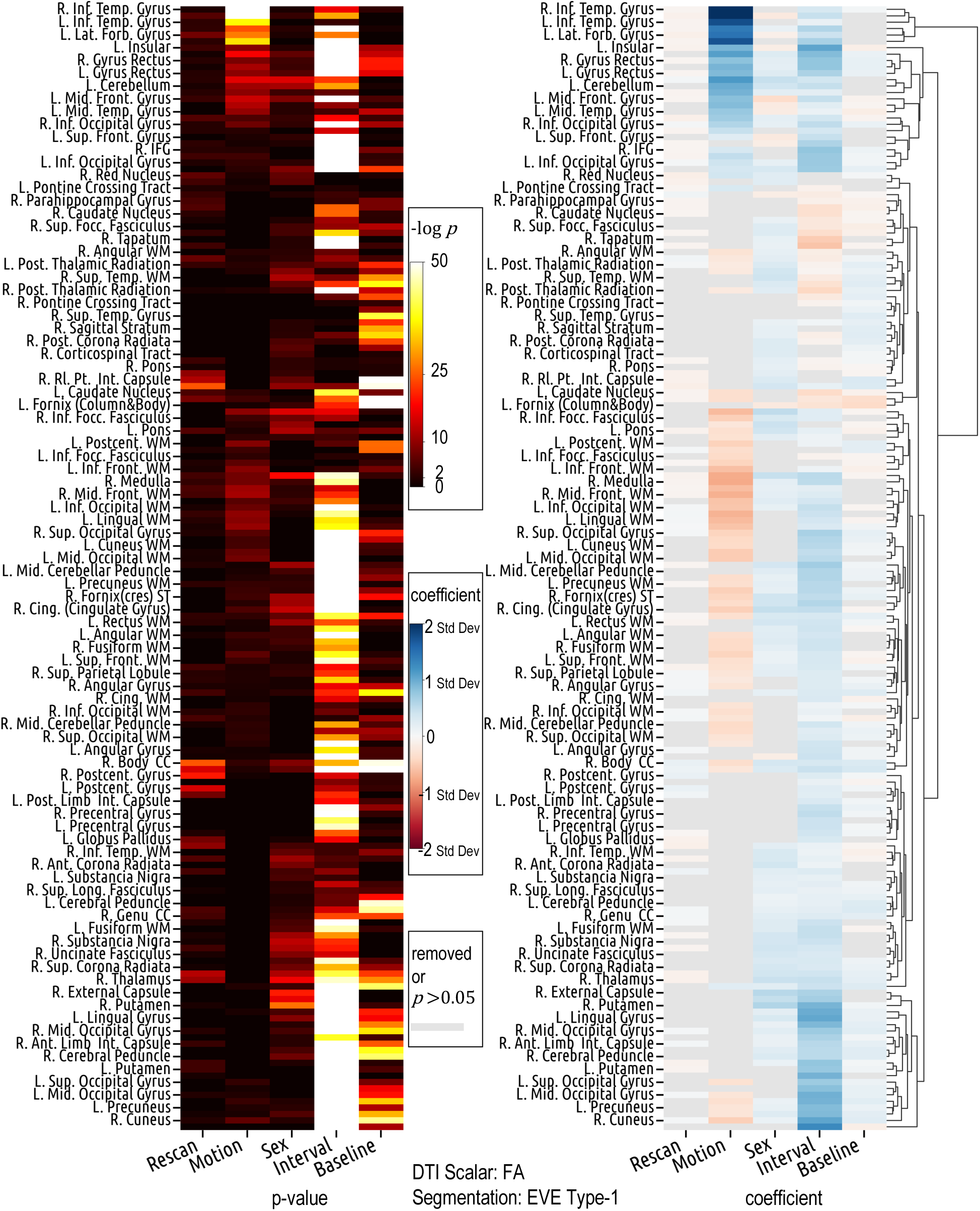
Covariate effects on FA standard deviation (standardized) are region-specific. Motion and interval exhibit opposite effect directions in many ROIs. Gender differences exist in multiple ROIs. Counterintuitively, motion is negatively related to FA standard deviation in many ROIs. The lookup table for the abbreviation of ROI name is in the Supplementary Materials.

**Table 2.** Covariate effects on FA standard deviation (standardized) in selected ROIs from Eve Type 1 atlas.

| Covariate<br>ROI | Age<sub>interval</sub> |  |  |  | Motion |  |  |  |
| --- | --- | --- | --- | --- | --- | --- | --- | --- |
|  | Left |  | Right |  | Left |  | Right |  |
| | $\beta$ | p-value | $\beta$ | p-value | $\beta$ | p-value | $\beta$ | p-value |
| Caudate Nucleus | -0.383 | 4.0e-41 | -0.317 | 2.4e-26 | -0.337 | 2.1e-03 | NA | NA |
| Posterior Thalamic Radiation | -0.091 | 1.1e-07 | -0.414 | 7.7e-94 | -0.186 | 5.0e-03 | -0.188 | 1.8e-02 |
| Superior Fronto-occipital Fasciculus | -0.358 | 2.5e-35 | -0.220 | 6.4e-11 | NA | NA | NA | NA |
| Middle Occipital Gyrus | 0.957 | 7.2e-218 | 0.726 | 1.1e-173 | -0.266 | 2.3e-02 | NA | NA |
| Lateral Fronto-orbital Gyrus | 0.427 | 5.5e-28 | 0.457 | 1.4e-31 | 1.481 | 3.6e-28 | 2.011 | 2.3e-51 |
| Insular | 1.055 | 1.7e-132 | 1.276 | 1.5e-208 | 0.935 | 5.8e-10 | NA | NA |
| Gyrus Rectus | 0.636 | 4.6e-59 | 0.787 | 1.2e-88 | 0.887 | 2.1e-10 | 0.818 | 5.2e-09 |
| Inferior Occipital Gyrus | 0.746 | 1.6e-109 | 0.505 | 2.6e-62 | 0.422 | 9.3e-4 | 0.597 | 3.4e-08 |
| Body of Corpus Callosum | 0.439 | 7.6e-60 | 0.336 | 2.6e-32 | -0.242 | 2.4e-02 | -0.357 | 1.1e-03 |
| Inferior Cerebellar Peduncle | 0.560 | 2.9e-55 | 0.605 | 1.7e-61 | -0.469 | 2.6e-04 | -0.346 | 1.1e-02 |
| Cingulum (Cingulate Gyrus) | 0.247 | 1.9e-17 | 0.498 | 3.0e-57 | -0.630 | 2.0e-10 | -0.476 | 1.2e-05 |
| Cuneus | 0.951 | 1.1e-288 | 1.061 | $\leq 2.3e-308$ | -0.318 | 3.2e-04 | -0.471 | 5.4e-07 |
| Cuneus WM | 0.555 | 2.0e-96 | 0.585 | 1.4e-95 | -0.383 | 4.0e-05 | -0.393 | 7.9e-05 |
| Fornix(cres) Stria Terminalis | 0.340 | 5.4e-41 | 0.509 | 9.0e-60 | -0.313 | 1.4e-03 | -0.433 | 9.9e-05 |
| Middle Frontal WM | 0.203 | 1.2e-15 | 0.275 | 5.4e-22 | -0.623 | 8.0e-12 | -0.716 | 2.5e-12 |
| Superior Frontal WM | 0.341 | 2.4e-48 | 0.338 | 3.5e-39 | -0.316 | 1.6e-04 | -0.440 | 7.3e-07 |
| Lateral Fronto-orbital WM | 0.372 | 7.3e-28 | 0.410 | 1.0e-33 | -0.566 | 1.7e-06 | -0.376 | 2.5e-03 |
| Lingual WM | 0.366 | 2.3e-36 | 0.426 | 6.1e-45 | -0.640 | 6.4e-10 | -0.695 | 1.5e-10 |
| Medulla | 0.537 | 1.9e-48 | 0.519 | 3.9e-45 | -0.744 | 1.6e-08 | -0.750 | 1.6e-08 |
| Superior Occipital Gyrus | 0.734 | 4.7e-163 | 0.644 | 1.3e-134 | -0.311 | 1.1e-03 | -0.445 | 1.7e-06 |
| Middle Occipital WM | 0.544 | 3.5e-102 | 0.308 | 2.3e-38 | -0.501 | 2.2e-08 | -0.564 | 3.5e-11 |
| Precuneus WM | 0.549 | 3.0e-96 | 0.499 | 1.7e-76 | -0.341 | 6.2e-04 | -0.508 | 1.4e-07 |

**Table 3.** Effects of sex on FA standard deviation (standardized) in selected ROIs from Eve Type 1 atlas.

| Covariate<br>ROI | Sex |  |  |  |
| --- | --- | --- | --- | --- |
|  | Left |  | Right |  |
| | $\beta$ | p-value | $\beta$ | p-value |
| Putamen | NA | NA | 0.602 | 2.6e-27 |
| Thalamus | 0.388 | 5.9e-13 | 0.450 | 4.3e-18 |
| Body of Corpus Callosum | 0.287 | 4.1e-07 | 0.353 | 1.3e-09 |
| Cingulum (Cingulate Gyrus) | 0.509 | 1.7e-16 | 0.471 | 1.4e-14 |
| Middle Frontal Gyrus | -0.370 | 1.8e-09 | -0.148 | 1.9e-02 |

In the lateral fronto-orbital gyrus, left insular, gyrus rectus, and inferior occipital gyrus, both motion and interval exhibit a positive association with FA variance (Table 2). In the left caudate nucleus, and right posterior thalamic radiation, they both show a negative association with FA variance (Table 2). In many other ROIs, such as the cuneus, lingual white matter, and middle occipital white matter, motion is negatively related to FA variance while interval is positively related (Table 2). Results from the left ROI closely align with those from the corresponding right ROI (Table 2). There are some ROIs where interval is significantly (*p* ≪ 0.001) associated with FA variance, while motion either gets removed during the model selection or shows weak associations (*p* ≥ 0.05) (Fig. 5).

On data extracted from ROIs defined by SLANT segmentation, which has different regional focus and delineation than Eve type-1 segmentation, the aforementioned patterns of effects can also be observed (Fig. 6, Fig. 7). For instance, in the left cerebellum exterior, both motion and interval are positively associated with FA variance, with motion’s coefficient (*β* = 0.960, *p* ≪ 0.001) higher than that of interval ( *β* = 0.485, *p* ≪ 0.001). This parallels the relationship observed between the motion and interval coefficients in the left cerebellum defined by the Eve type-1 segmentation ( *β* = 0.993, *p* ≪ 0.001 for motion; *β* = 0.471, *p* ≪ 0.001 for interval). Similarly, in ROIs such as the right cuneus, left precuneus, right superior occipital gyrus, and right middle occipital gyrus, motion shows a negative association with FA variance, while interval shows a positive association. The exact coefficients and p-values are in the supplementary materials, where we also provide the results from other pairs of DTI scalar and segmentation method.

**Fig. 6.**
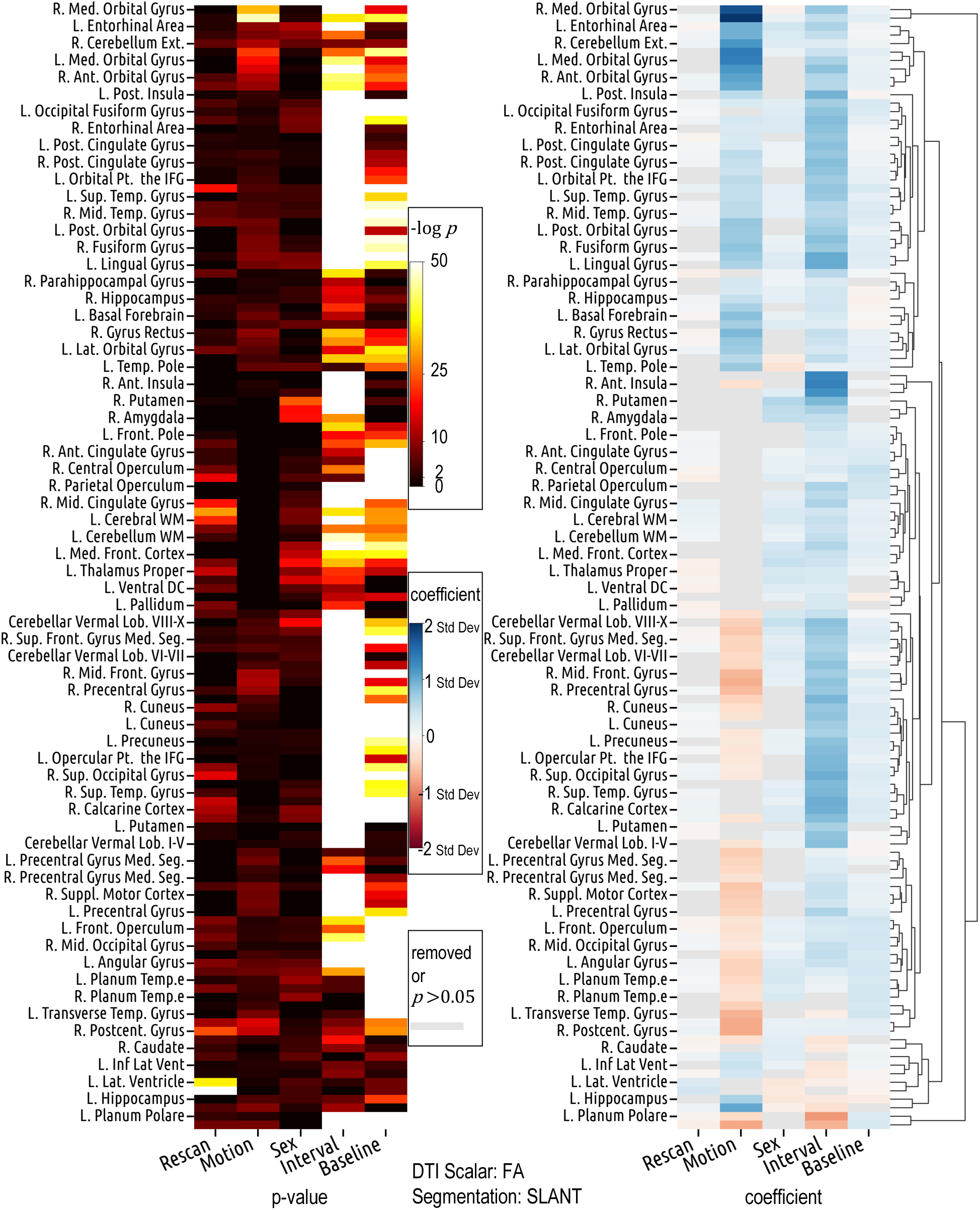
The region-specific and bidirectional patterns of covariate effects are similarly observed in the results derived from SLANT segmentation, despite its differing definitions and delineations of ROIs compared to Eve type-1 segmentation (Fig. 5).

**Fig. 7.**
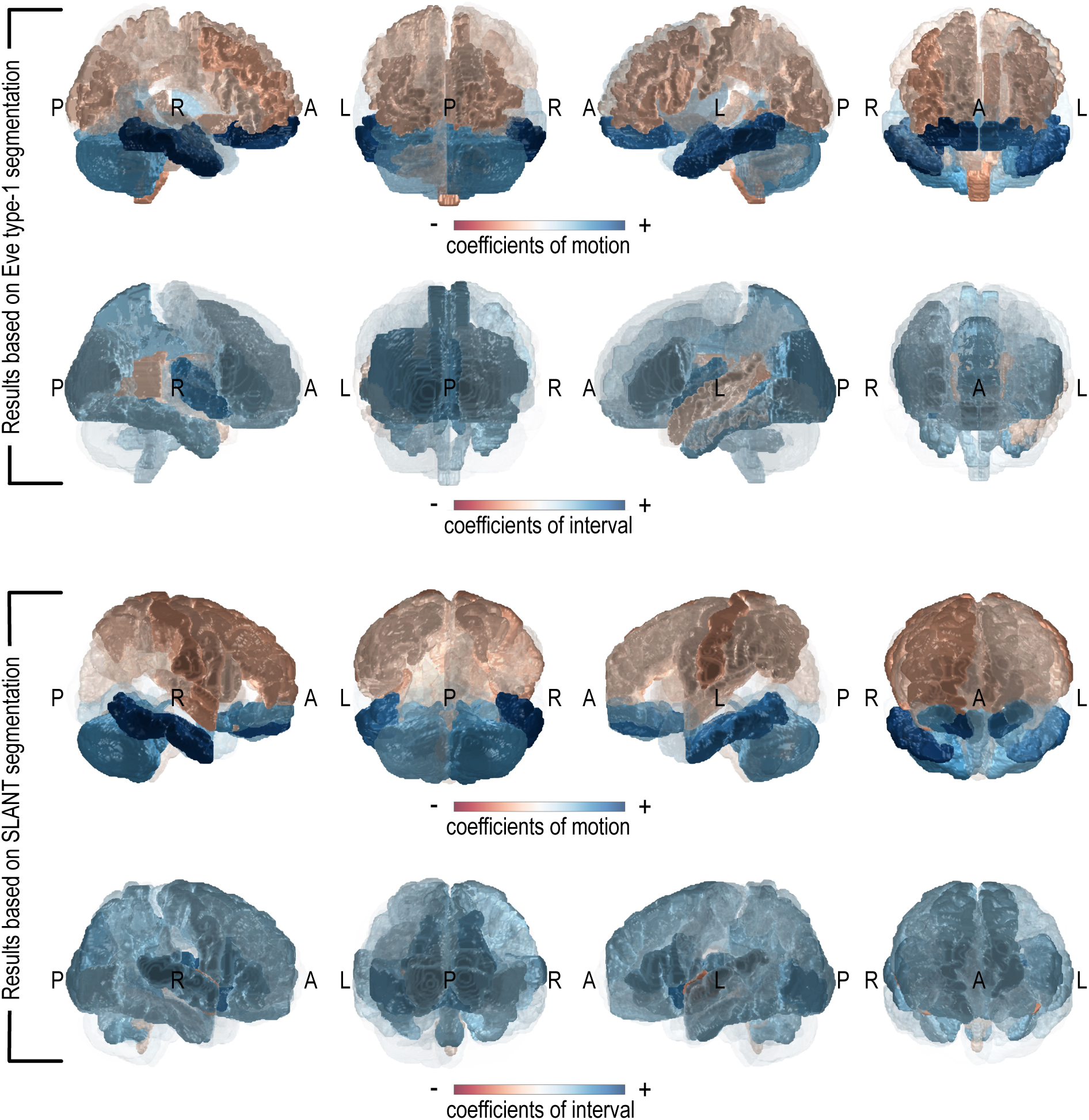
Despite the different definitions and delineations of ROIs between Eve type-1 and SLANT segmentations, results based on the two segmentation methods are largely similar (comparable regions are colored similarly) and both show that the effects of motion and interval on FA variance vary across ROIs.

In the model selection process, we observe that the rescan and the motion terms appear mutually exclusive, with only one preserved post-selection in most models. This pattern is echoed in the heatmaps of coefficients (Fig. 5, Fig. 6), where the cell in either the motion or the rescan column is colored grey. This hints at a correlation between rescan and motion. Supporting this observation, we detect an increase in head motion in the rescan of DTI acquired right after the first scan of DTI in the same session (mean shift Δ*μ* = 0.045 millimeters per volume, relative mean shift Δ*μ*/*μ* = 17.0%, coefficient of determination *R*^2^ = 0.065).

To examine the differences between scanners, we measure the signal-to-noise ratio (SNR) of the FA images based on ROIs. There are differences between scanners regarding SNR across ROIs (Fig. 8). To validate our findings, we include two additional datasets, ADNI^40^ and BIOCARD^41^. We exclude data points from subjects with cognitive impairment and use the remaining 1808 subjects from the three datasets for the experiment. Detailed information about the data is included in the supplementary materials. We reproduce the experiments of the linear mixed-effects models, except that the rescan term in equation (4) is omitted because the definitions of rescan in ADNI and BIOCARD differ from the definition of rescan in BLSA. The coefficients and p-values, visualized in Fig. 9, show a similar pattern to those derived from the BLSA, although the effect sizes differ, and the hierarchical clustering differs partly due to the omission of one covariate.

**Fig. 8.**
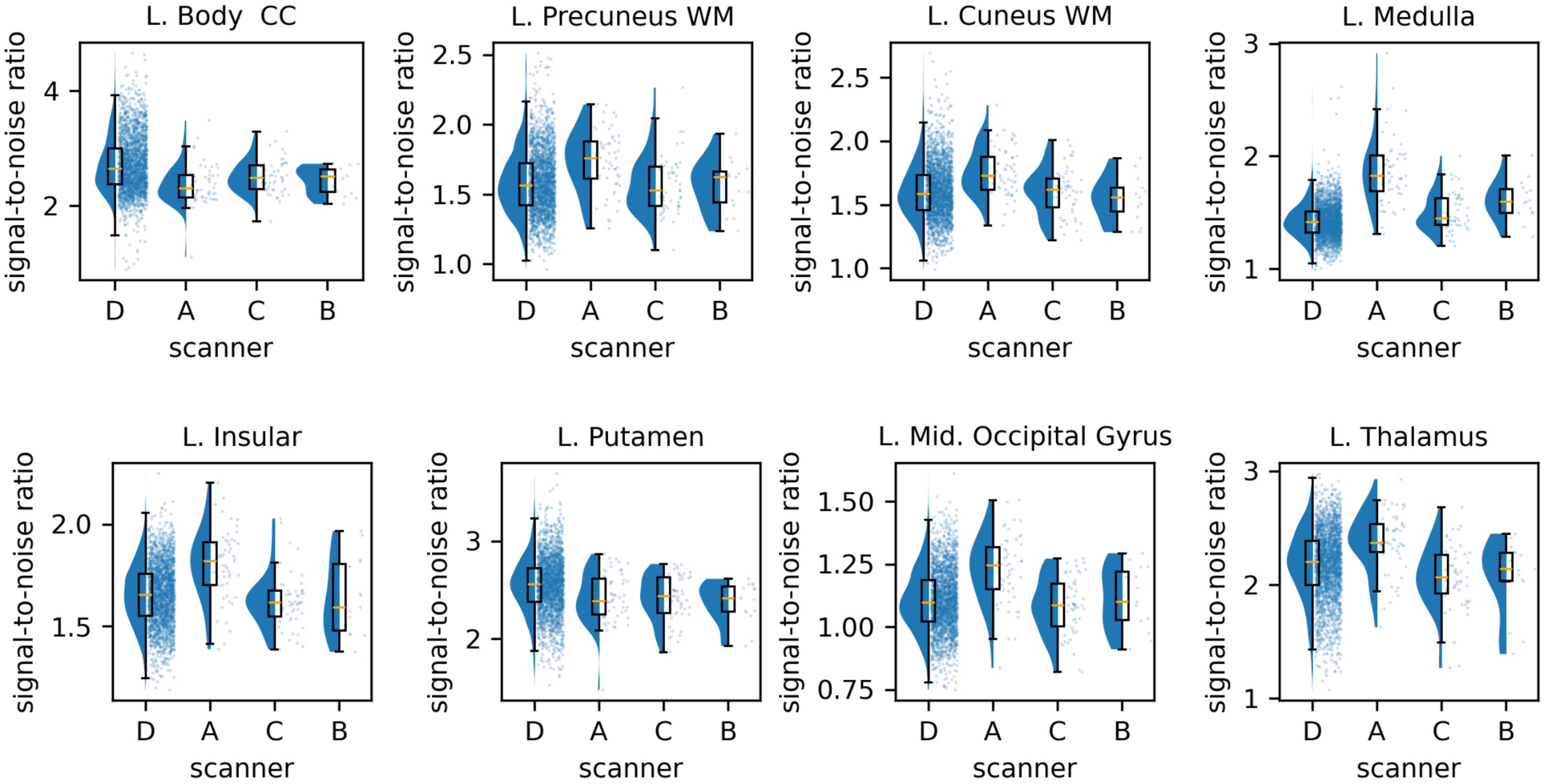
Signal-to-noise ratio (SNR) of the FA images across 8 Eve type-1 atlas-defined ROIs, including white matter (WM) regions (body of the corpus callosum, precuneus WM, cuneus WM), gray matter regions (insular, putamen, middle occipital gyrus, thalamus), and mixed regions (medulla), in 4 different scanners of BLSA (where scanner A is the 1.5 Tesla Philips Intera scanner, and scanners B/C/D are the 3 Tesla Philips Achieva scanners). “L.” stands for the left hemisphere of the brain.

**Fig. 9.**
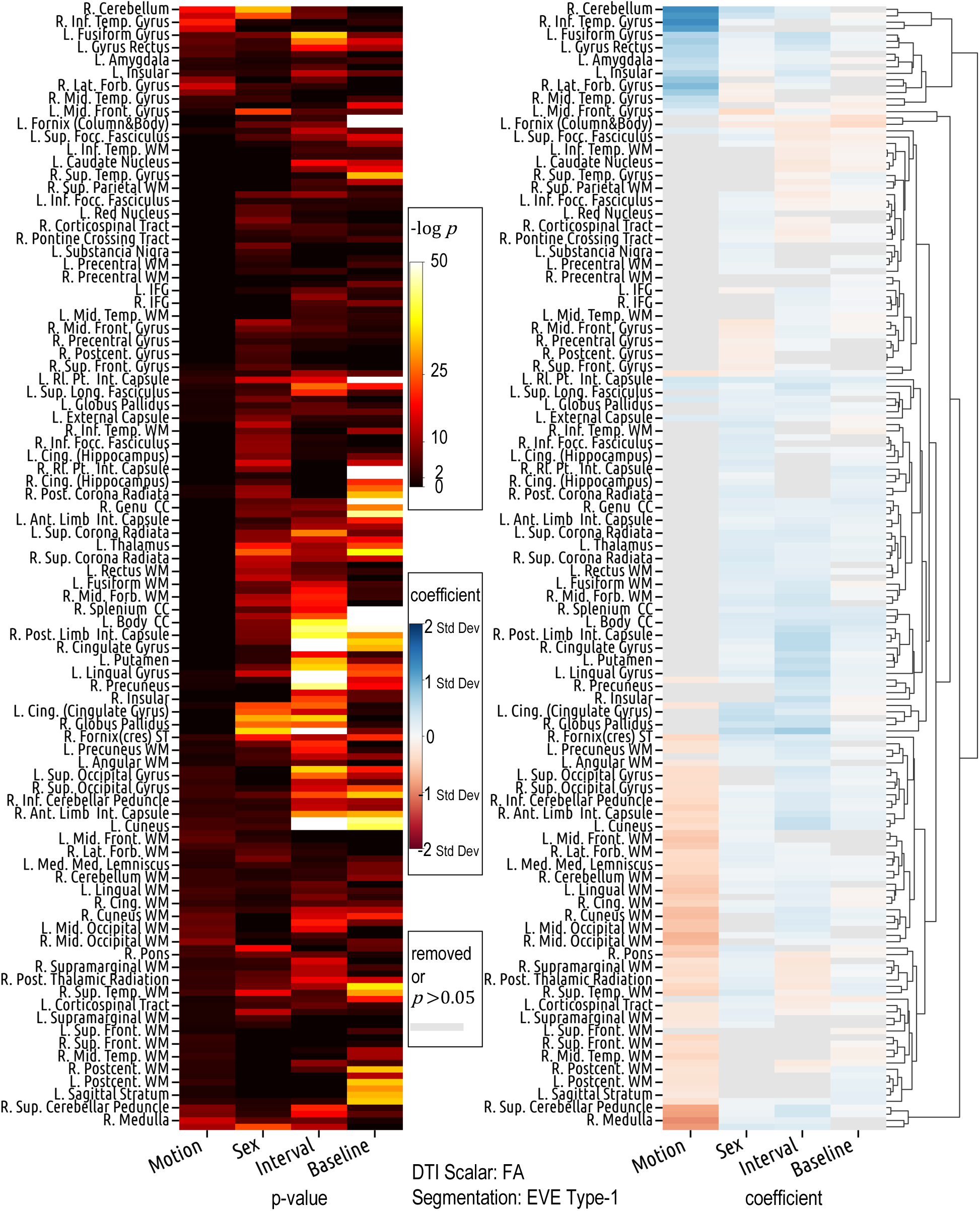
To assess the generalizability of our findings, we include two additional datasets, ADNI and BIOCARD and fit the linear mixed-effects models. The coefficients and p-values show similar patterns of those from BLSA alone, despite that the effect sizes and the hierarchical clustering are different, partly due to the omission of the rescan covariate.

## 4 Discussion

While many studies have estimated and shown the spatial variability of DTI variance (or noise),^14,15,42^ we characterize how DTI variance is associated with physiological and behavioral factors across brain regions. We answer the questions: Which factors are associated with DTI variance? Where and how does this association manifest? We found region-specific and bidirectional effects of covariates—including interval (which captures the within-individual longitudinal change over time), motion, and sex—on FA variance across brain regions. For instance, FA variance is positively associated with interval in cuneus, but negatively associated in caudate nucleus. Long-standing research has demonstrated that there is a decline in white matter microstructure with aging,^43–48^ with the consensus being that frontal and parietal areas are particularly vulnerable and the occipital and motor areas are mostly preserved. The frontal lobe exhibits the most pronounced decline, with FA declining by approximately 3% per decade starting at ∼35 years of age.^49^ Although our study focuses on the standard deviation of FA, our results converge with these prior research studies as we have shown high sensitivity to aging in the frontal, parietal, and temporal areas. While it is unclear what mechanisms are driving the changes in these areas, potential culprits include the change of uniformity of fiber orientations and fiber density.^50–52^

Previous studies^53–55^ have shown differences in FA between genders across brain regions. Oh et al. found that males have significantly higher FA values in global corpus callosum structure areas, while they exhibit lower FA values than females in the partial areas of the rostrum, genu, and splenium.^53^ Menzler et al. found that males show higher FA values in the thalamus, corpus callosum and cingulum.^55^ Most of these regions previously identified in the literature also show significant (*p* ≪ 0.001) associations between FA variance and sex in our study. While previous studies have reported changes in mean FA values, we offer a different perspective by depicting the variance of FA values.

The negative association observed between motion and FA variance in multiple regions, while counterintuitive, is not unreasonable. One might naturally expect that as motion increases, the uncertainty (reflected as variance) in the image should increase, given that motion leads to lower image quality, signal-to-noise ratio, and artifacts that can mislead image interpretation.^56,57^ However, the images we use for analysis have undergone motion correction during preprocessing. Although in practice, motion artifacts cannot be fully eliminated from the image, the recorded motion value doesn’t reflect the motion’s impact in the image after preprocessing. Instead, it reflects the subject’s motion during image acquisition. A higher motion value does not necessarily correspond to a noisier image post-preprocessing. Furthermore, Zeng et al. found that head motion during brain imaging is not merely a technical artifact but a reflection of a neurobiological trait. Specifically, individuals with stronger distant connectivity in the default network could consistently refrain from moving and such “head motion tendency” remains consistent within individuals.^58^ These points, taken together, provide explanations from image processing and biological perspectives, respectively, for why FA variance can decrease as motion increases.

This study underscores the significance of heteroscedasticity in diffusion-weighted MRI mega-analyses and provides a relatively straightforward approach to addressing this issue. Despite the longstanding recognition of heteroscedasticity in statistical analyses,^59–62^ its application to diffusion-weighted MRI is still in its nascent stages. Recent advancements have started to bridge this gap, with emerging studies illustrating the importance of accounting for heteroscedasticity in MRI data.^63–65^ These pioneering efforts are pivotal, yet they remain underutilized in the broader research community. To enhance the precision and reliability of findings in mega-analyses, it is imperative to disseminate these methodologies more widely and integrate them into commonly used analytical tools. Future research should focus on developing and employing increasingly sophisticated techniques to model and understand heteroscedasticity, thereby improving the robustness of statistical assessments in large-scale neuroimaging studies.

### 4.1 Limitations of Current Study

First, this study relies on a registration-based method for brain segmentation in the b0 space. Despite rigorous quality assurance, the labels for each brain region may not correspond flawlessly with the true anatomical regions. Consequently, the standard deviation of DTI scalars extracted from each region combines both voxel-wise modeling factors and image analysis factors from neighboring regions. Second, we used backward model selection for the fixed-effect terms of the linear mixed-effects models. Such method can be unstable according to Breiman et. al.^66^ Third, we use the variance of DTI scalar values in the ROI as a proxy for measuring noise. This is not an ideal proxy, because signal intensities may not be homogeneously distributed within each ROI, and the ROI-based variance captures not only the image noise but also the spatial variability in voxel intensity due to microstructural variations. This makes it suboptimal to reflect DTI noise. Fourth, the motion value used in this study is based on movement calculated by FSL’s eddy^38^, which approximates true head motion. Additional sensors or motion tracking sequences might be necessary to quantify head motion during scanning more accurately.

## 5 Conclusion

The notion of harnessing variance to enhance the reliability of analysis is universally applicable. Having a better understanding of variance is pivotal in mega-analyses, where heteroscedasticity is often an inherent challenge. Our study illuminates the complex and heterogeneous effects of covariates including baseline age, interval, motion, sex, and rescan on DTI variance across ROIs. More comprehensive efforts are required to fully characterize the variance. In the meantime, we encourage researchers to consider models of heteroscedasticity in their analyses and to include their estimates of variance when sharing data. As highlighted in the introduction, the application of the whitening matrix, constructed using the variance of the data, significantly reduces statistical errors. We anticipate that more sophisticated methods can further unlock the potential benefits derived from a nuanced understanding of variance, thereby bolstering the accuracy and reliability of future research.

## Supporting information

Supplemental Table - LME Stats (SLANT - FA)

Supplemental Table - LME Stats (EveType1 - FA)

Supplemental Table - Lookup Table for ROIs

Supplemental Figures - Part 1

Supplemental Figures - Part 2

## Data Availability

All data used in the present work are available upon request to the Baltimore Longitudinal Study of Aging (BLSA), the Alzheimer's Disease Neuroimaging Initiative (ADNI), and the BIOCARD study.

https://www.blsa.nih.gov/

https://www.biocard-se.org/public/BIOCARD%20Home%20Page.html

https://adni.loni.usc.edu/

## Code, Data, and Materials Availability

The dataset used in this study can be accessed upon approval from the Baltimore Longitudinal Study of Aging team at https://www.blsa.nih.gov/. The code for the experiments can be found on GitHub at https://github.com/MASILab/Variance-Aging-Diffusion.

## Acknowledgements

This work was supported in part by the National Institute of Health through grants 1R01EB017230-01A1 (Bennett A. Landman), 5-K01-EB032898-02 (Kurt Schilling), 1U24AG074855-01 (Timothy J. Hohman), and ViSE/VICTR VR3029 and by the Intramural Research Program of the National Institute on Aging, NIH. This work was conducted in part using the resources of the Advanced Computing Center for Research and Education at Vanderbilt University, Nashville, TN. We appreciate the National Institute of Health S10 Shared Instrumentation grant 1S10OD020154-01 (Smith), and grant 1S10OD023680-01 (Vanderbilt’s High-Performance Computer Cluster for Biomedical Research). The Vanderbilt Institute for Clinical and Translational Research (VICTR) is funded by the National Center for Advancing Translational Sciences (NCATS) Clinical Translational Science Award (CTSA) Program, Award Number 5UL1TR002243-03. The content is solely the responsibility of the authors and does not necessarily represent the official views of the NIH.

The BLSA is supported by the Intramural Research Program, National Institute on Aging, NIH. The BIOCARD study is supported by a grant from the National Institute on Aging (NIA): U19- AG03365. The BIOCARD Study consists of 7 Cores and 2 projects with the following members: (1) The Administrative Core (Marilyn Albert, Corinne Pettigrew, Barbara Rodzon); (2) the Clinical Core (Marilyn Albert, Anja Soldan, Rebecca Gottesman, Corinne Pettigrew, Leonie Farrington, Maura Grega, Gay Rudow, Rostislav Brichko, Scott Rudow, Jules Giles, Ned Sacktor); (3) the Imaging Core (Michael Miller, Susumu Mori, Anthony Kolasny, Hanzhang Lu, Kenichi Oishi, Tilak Ratnanather, Peter vanZijl, Laurent Younes); (4) the Biospecimen Core (Abhay Moghekar, Jacqueline Darrow, Alexandria Lewis, Richard O’Brien); (5) the Informatics Core (Roberta Scherer, Ann Ervin, David Shade, Jennifer Jones, Hamadou Coulibaly, Kathy Moser, Courtney Potter); the (6) Biostatistics Core (Mei-Cheng Wang, Yuxin Zhu, Jiangxia Wang); (7) the Neuropathology Core (Juan Troncoso, David Nauen, Olga Pletnikova, Karen Fisher); (8) Project 1 (Paul Worley, Jeremy Walston, Mei-Fang Xiao), and (9) Project 2 (Mei-Cheng Wang, Yifei Sun, Yanxun Xu. Data collection and sharing for ADNI were supported by National Institutes of Health Grant U01-AG024904 and Department of Defense (award number W81XWH- 12-2-0012). ADNI is also funded by the National Institute on Aging, the National Institute of Biomedical Imaging and Bioengineering, and through generous contributions from the following: AbbVie, Alzheimer’s Association; Alzheimer’s Drug Discovery Foundation; Araclon Biotech; BioClinica, Inc.; Biogen; Bristol-Myers Squibb Company; CereSpir, Inc.; Cogstate; Eisai Inc.; Elan Pharmaceuticals, Inc.; Eli Lilly and Company; EuroImmun; F. Hoffmann-La Roche Ltd and its affiliated company Genentech, Inc.; Fujirebio; GE Healthcare; IXICO Ltd.; Janssen Alzheimer Immunotherapy Research & Development, LLC.; Johnson & Johnson Pharmaceutical Research & Development LLC.; Lumosity; Lundbeck; Merck & Co., Inc.; Meso Scale Diagnostics, LLC.; NeuroRx Research; Neurotrack Technologies; Novartis Pharmaceuticals Corporation; Pfizer Inc.; Piramal Imaging; Servier; Takeda Pharmaceutical Company; and Transition Therapeutics. The Canadian Institutes of Health Research is providing funds to support ADNI clinical sites in Canada. Private sector contributions are facilitated by the Foundation for the National Institutes of Health (www.fnih.org). The grantee organization is the Northern California Institute for Research and Education, and the study is coordinated by the Alzheimer’s Therapeutic Research Institute at the University of Southern California. ADNI data are disseminated by the Laboratory for Neuro Imaging at the University of Southern California.

## Ethics approval

IRB of Vanderbilt University waived ethical approval for de-identified access of the human subject data. (IRB #172072, PI: Bennett A. Landman)

## Declaration of Generative AI and AI-assisted technologies in the writing process

During the preparation of this work the author Chenyu Gao used ChatGPT to check grammar and improve readability of the original draft. After using this tool, the author reviewed and edited the content as needed and take full responsibility for the content of the publication.

**Chenyu Gao** is a Ph.D. student in electrical and computer engineering at Vanderbilt University, working with Prof. Bennett Landman on the harmonization of diffusion MRI. His research interest is focused on image processing and computer vision with application to medical image analysis. He received his BS degree in biomedical engineering from Sun Yat-sen University, and his MS degree in biomedical engineering from Johns Hopkins University, working with Prof. Jerry Prince.

