## Supplemental Figures - Part 1 for "Characterizing patterns of diffusion tensor imaging variance in aging brains"

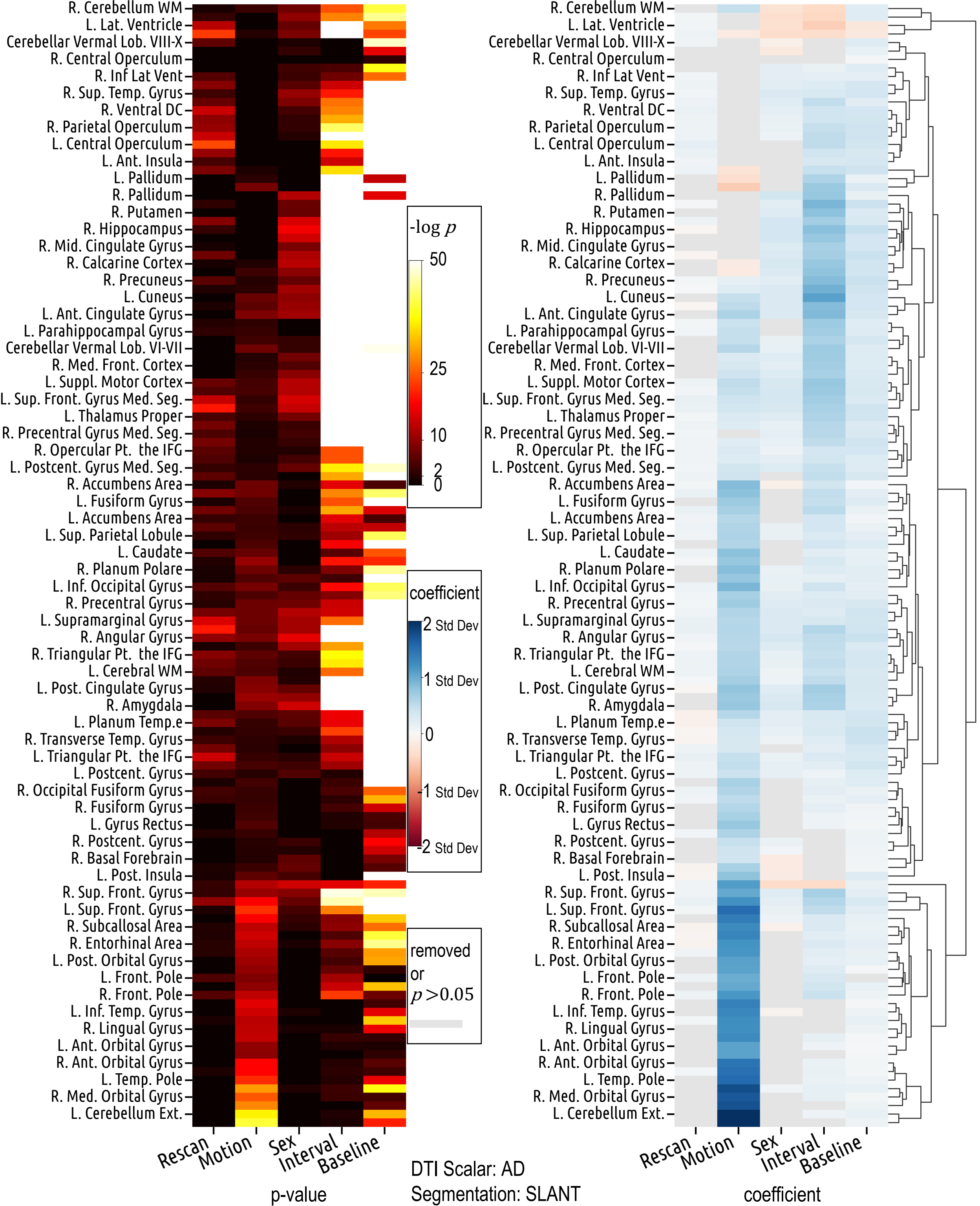

R. Cerebellum WM
L. Lat. Ventricle
Cerebellar Vermal Lob. VIII-X
R. Central Operculum
R. Inf Lat Vent
R. Sup. Temp. Gyrus
R. Ventral DC
R. Parietal Operculum
L. Central Operculum
L. Ant. Insula
L. Pallidum
R. Pallidum
R. Putamen
R. Hippocampus
R. Mid. Cingulate Gyrus
R. Calcarine Cortex
R. Precuneus
L. Cuneus
L. Ant. Cingulate Gyrus
L. Parahippocampal Gyrus
Cerebellar Vermal Lob. VI-VII
R. Med. Front. Cortex
L. Suppl. Motor Cortex
L. Sup. Front. Gyrus Med. Seg.
L. Thalamus Proper
R. Precentral Gyrus Med. Seg.
R. Opercular Pt. the IFG
L. Postcent. Gyrus Med. Seg.
R. Accumbens Area
L. Fusiform Gyrus
L. Accumbens Area
L. Sup. Parietal Lobule
L. Caudate
R. Planum Polare
L. Inf. Occipital Gyrus
R. Precentral Gyrus
L. Supramarginal Gyrus
R. Angular Gyrus
R. Triangular Pt. the IFG
L. Cerebral WM
L. Post. Cingulate Gyrus
R. Amygdala
L. Planum Temp.e
R. Transverse Temp. Gyrus
L. Triangular Pt. the IFG
L. Postcent. Gyrus
R. Occipital Fusiform Gyrus
R. Fusiform Gyrus
L. Gyrus Rectus
R. Postcent. Gyrus
R. Basal Forebrain
L. Post. Insula
R. Sup. Front. Gyrus
L. Sup. Front. Gyrus
R. Subcallosal Area
R. Entorhinal Area
L. Post. Orbital Gyrus
L. Front. Pole
R. Front. Pole
L. Inf. Temp. Gyrus
R. Lingual Gyrus
L. Ant. Orbital Gyrus
R. Ant. Orbital Gyrus
L. Temp. Pole
R. Med. Orbital Gyrus
L. Cerebellum Ext.
-log p
50
25
10
2
0
coefficient
2 Std Dev
1 Std Dev
0
-1 Std Dev
-2 Std Dev
removed or p>0.05
Rescan
Motion
Sex
Interval
Baseline
p-value
DTI Scalar: AD
Segmentation: SLANT
coefficient

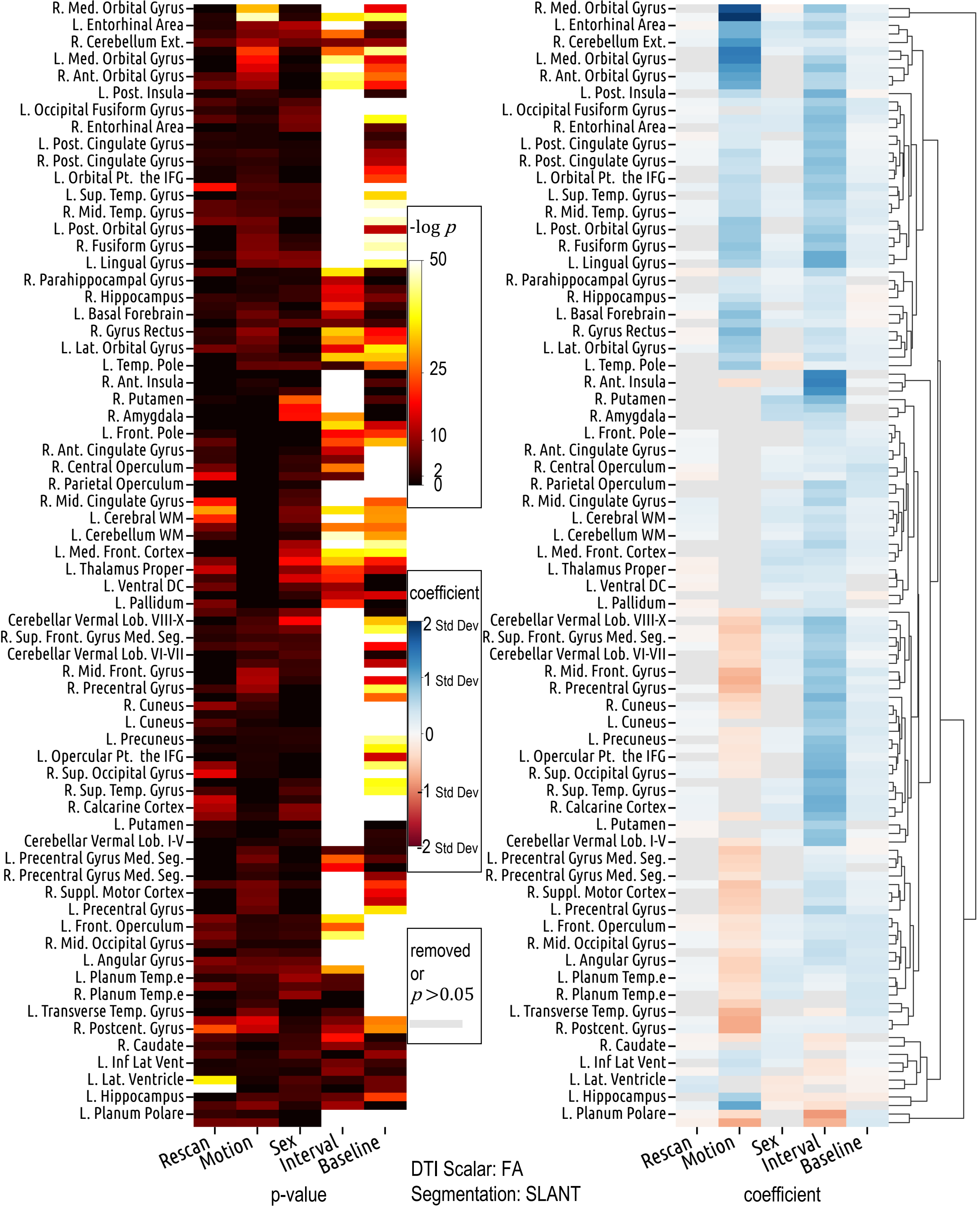

R. Med. Orbital Gyrus
L. Entorhinal Area
R. Cerebellum Ext.
L. Med. Orbital Gyrus
R. Ant. Orbital Gyrus
L. Post. Insula
L. Occipital Fusiform Gyrus
R. Entorhinal Area
L. Post. Cingulate Gyrus
R. Post. Cingulate Gyrus
L. Orbital Pt. the IFG
L. Sup. Temp. Gyrus
R. Mid. Temp. Gyrus
L. Post. Orbital Gyrus
R. Fusiform Gyrus
L. Lingual Gyrus
R. Parahippocampal Gyrus
R. Hippocampus
L. Basal Forebrain
R. Gyrus Rectus
L. Lat. Orbital Gyrus
L. Temp. Pole
R. Ant. Insula
R. Putamen
R. Amygdala
L. Front. Pole
R. Ant. Cingulate Gyrus
R. Central Operculum
R. Parietal Operculum
R. Mid. Cingulate Gyrus
L. Cerebral WM
L. Cerebellum WM
L. Med. Front. Cortex
L. Thalamus Proper
L. Ventral DC
L. Pallidum
Cerebellar Vermal Lob. VIII-X
R. Sup. Front. Gyrus Med. Seg.
Cerebellar Vermal Lob. VI-VII
R. Mid. Front. Gyrus
R. Precentral Gyrus
R. Cuneus
L. Cuneus
L. Precuneus
L. Opercular Pt. the IFG
R. Sup. Occipital Gyrus
R. Sup. Temp. Gyrus
R. Calcarine Cortex
L. Putamen
Cerebellar Vermal Lob. I-V
L. Precentral Gyrus Med. Seg.
R. Precentral Gyrus Med. Seg.
R. Suppl. Motor Cortex
L. Precentral Gyrus
L. Front. Operculum
R. Mid. Occipital Gyrus
L. Angular Gyrus
L. Planum Temp.e
R. Planum Temp.e
L. Transverse Temp. Gyrus
R. Postcent. Gyrus
R. Caudate
L. Inf Lat Vent
L. Lat. Ventricle
L. Hippocampus
L. Planum Polare
Rescan
Motion
Sex
Interval
Baseline
p-value
-log p
50
25
10
2
0
coefficient
2 Std Dev
1 Std Dev
0
-1 Std Dev
-2 Std Dev
removed
or
p >0.05
DTI Scalar: FA
Segmentation: SLANT
coefficient

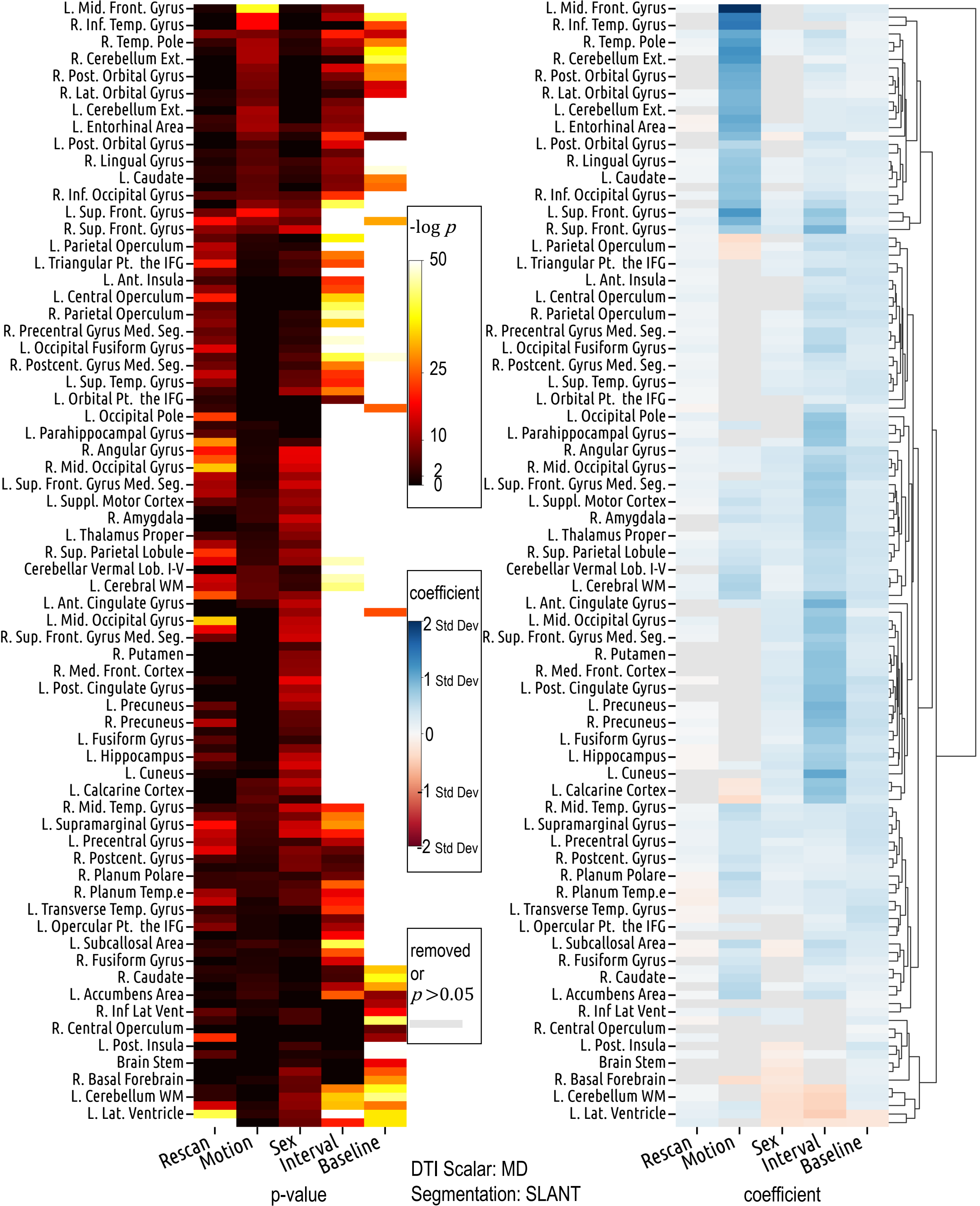

L. Mid. Front. Gyrus
R. Inf. Temp. Gyrus
R. Temp. Pole
R. Cerebellum Ext.
R. Post. Orbital Gyrus
R. Lat. Orbital Gyrus
L. Cerebellum Ext.
L. Entorhinal Area
L. Post. Orbital Gyrus
R. Lingual Gyrus
L. Caudate
R. Inf. Occipital Gyrus
L. Sup. Front. Gyrus
R. Sup. Front. Gyrus
L. Parietal Operculum
L. Triangular Pt. the IFG
L. Ant. Insula
L. Central Operculum
R. Parietal Operculum
R. Precentral Gyrus Med. Seg.
L. Occipital Fusiform Gyrus
R. Postcent. Gyrus Med. Seg.
L. Sup. Temp. Gyrus
L. Orbital Pt. the IFG
L. Occipital Pole
L. Parahippocampal Gyrus
R. Angular Gyrus
R. Mid. Occipital Gyrus
L. Sup. Front. Gyrus Med. Seg.
L. Suppl. Motor Cortex
R. Amygdala
L. Thalamus Proper
R. Sup. Parietal Lobule
Cerebellar Vermal Lob. I-V
L. Cerebral WM
L. Ant. Cingulate Gyrus
L. Mid. Occipital Gyrus
R. Sup. Front. Gyrus Med. Seg.
R. Putamen
R. Med. Front. Cortex
L. Post. Cingulate Gyrus
L. Precuneus
R. Precuneus
L. Fusiform Gyrus
L. Hippocampus
L. Cuneus
L. Calcarine Cortex
R. Mid. Temp. Gyrus
L. Supramarginal Gyrus
L. Precentral Gyrus
R. Postcent. Gyrus
R. Planum Polare
R. Planum Temp.e
L. Transverse Temp. Gyrus
L. Opercular Pt. the IFG
L. Subcallosal Area
R. Fusiform Gyrus
R. Caudate
L. Accumbens Area
R. Inf Lat Vent
R. Central Operculum
L. Post. Insula
Brain Stem
R. Basal Forebrain
L. Cerebellum WM
L. Lat. Ventricle
-log p
50
25
10
2
0
coefficient
2 Std Dev
1 Std Dev
0
-1 Std Dev
-2 Std Dev
removed
or
p>0.05
Rescan
Motion
Sex
Interval
Baseline
p-value
DTI Scalar: MD
Segmentation: SLANT
coefficient

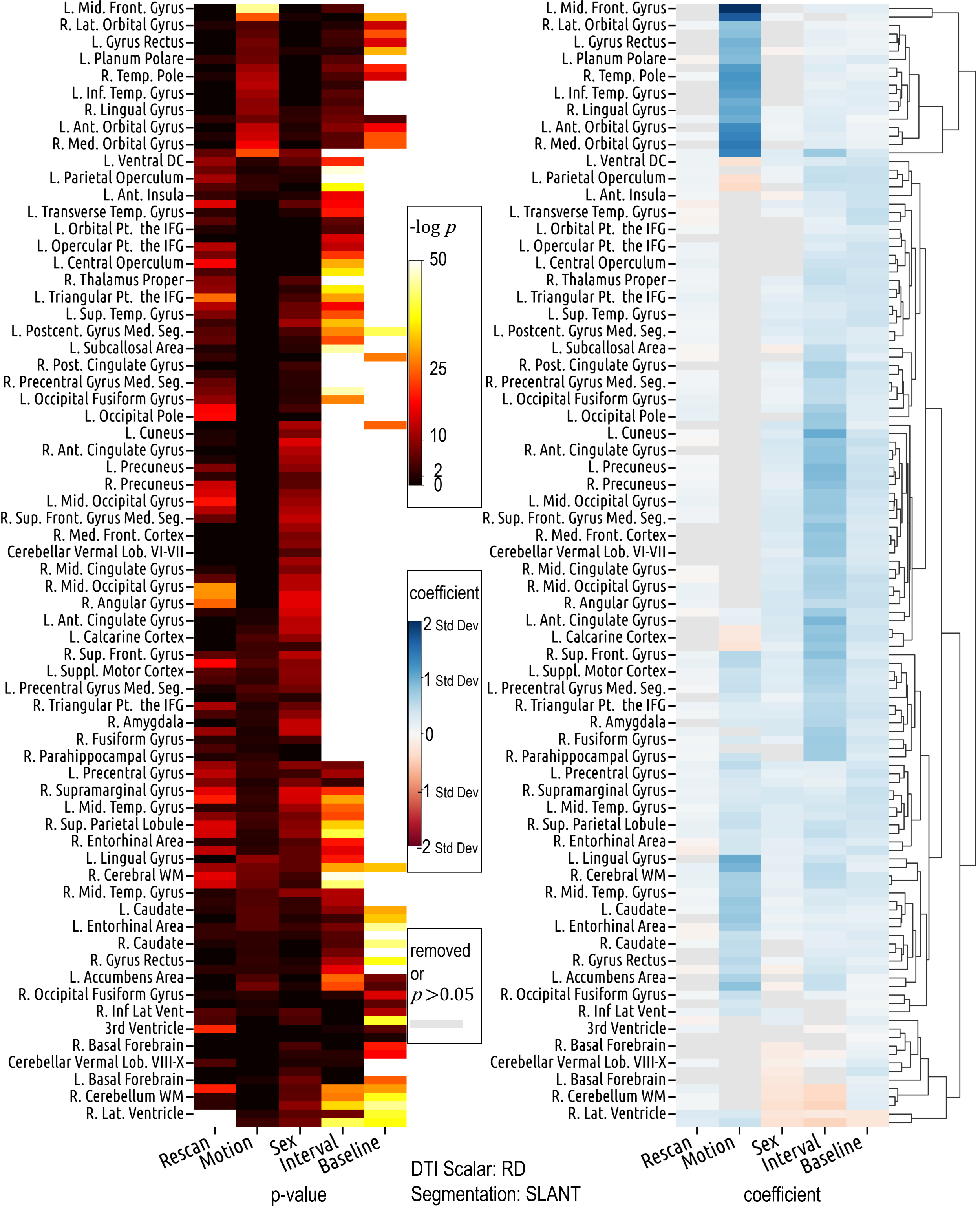

L. Mid. Front. Gyrus
R. Lat. Orbital Gyrus
L. Gyrus Rectus
L. Planum Polare
R. Temp. Pole
L. Inf. Temp. Gyrus
R. Lingual Gyrus
L. Ant. Orbital Gyrus
R. Med. Orbital Gyrus
L. Ventral DC
L. Parietal Operculum
L. Ant. Insula
L. Transverse Temp. Gyrus
L. Orbital Pt. the IFG
L. Opercular Pt. the IFG
L. Central Operculum
R. Thalamus Proper
L. Triangular Pt. the IFG
L. Sup. Temp. Gyrus
L. Postcent. Gyrus Med. Seg.
L. Subcallosal Area
R. Post. Cingulate Gyrus
R. Precentral Gyrus Med. Seg.
L. Occipital Fusiform Gyrus
L. Occipital Pole
L. Cuneus
R. Ant. Cingulate Gyrus
L. Precuneus
R. Precuneus
L. Mid. Occipital Gyrus
R. Sup. Front. Gyrus Med. Seg.
R. Med. Front. Cortex
Cerebellar Vermal Lob. VI-VII
R. Mid. Cingulate Gyrus
R. Mid. Occipital Gyrus
R. Angular Gyrus
L. Ant. Cingulate Gyrus
L. Calcarine Cortex
R. Sup. Front. Gyrus
L. Suppl. Motor Cortex
L. Precentral Gyrus Med. Seg.
R. Triangular Pt. the IFG
R. Amygdala
R. Fusiform Gyrus
R. Parahippocampal Gyrus
L. Precentral Gyrus
R. Supramarginal Gyrus
L. Mid. Temp. Gyrus
R. Sup. Parietal Lobule
R. Entorhinal Area
L. Lingual Gyrus
R. Cerebral WM
R. Mid. Temp. Gyrus
L. Caudate
L. Entorhinal Area
R. Caudate
R. Gyrus Rectus
L. Accumbens Area
R. Occipital Fusiform Gyrus
R. Inf Lat Vent
3rd Ventricle
R. Basal Forebrain
Cerebellar Vermal Lob. VIII-X
L. Basal Forebrain
R. Cerebellum WM
R. Lat. Ventricle
Rescan
Motion
Sex
Interval
Baseline
p-value
-log p
50
25
10
2
0
coefficient
2 Std Dev
1 Std Dev
0
-1 Std Dev
-2 Std Dev
removed or $p > 0.05$
DTI Scalar: RD
Segmentation: SLANT
Rescan
Motion
Sex
Interval
Baseline
coefficient

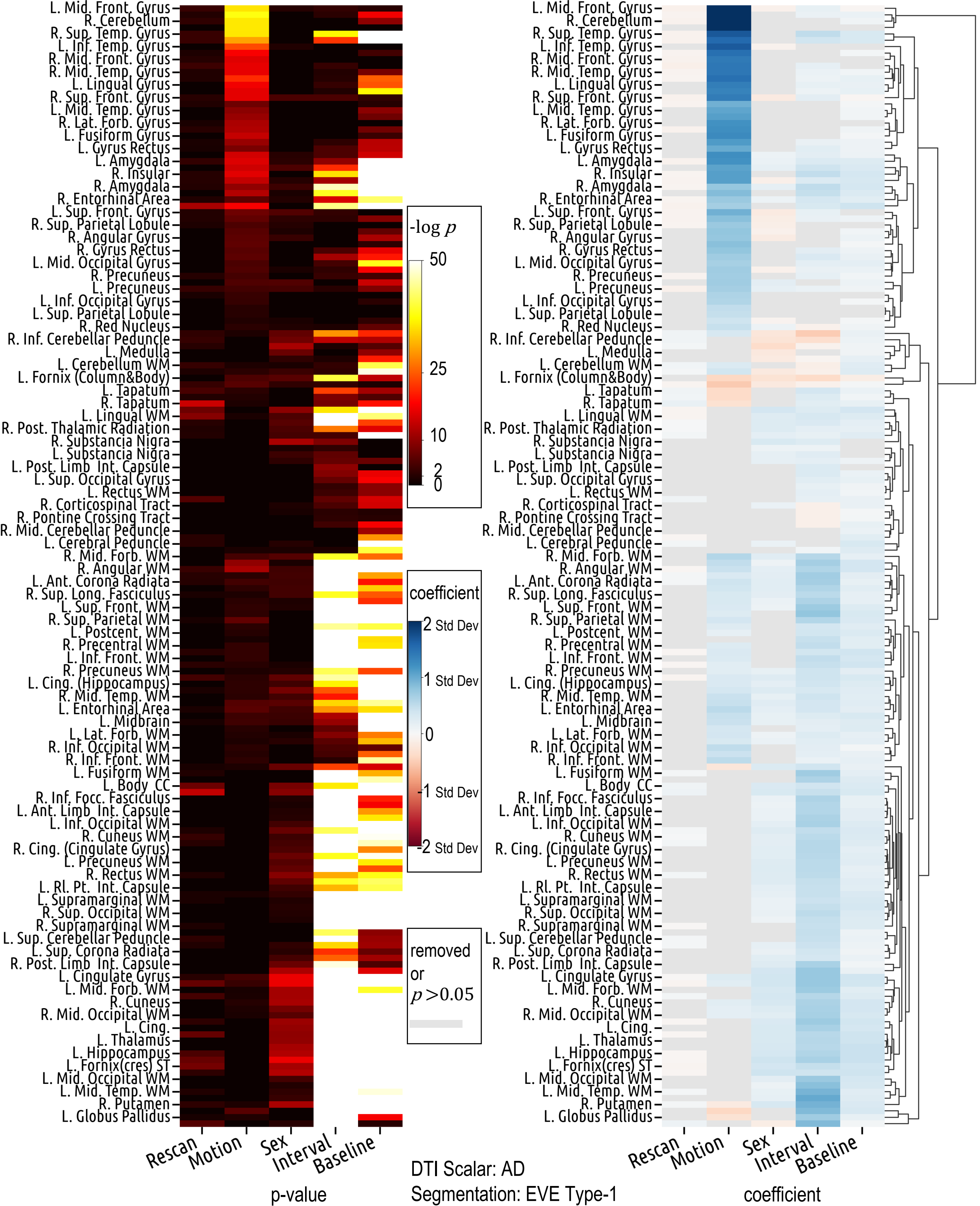

L. Mid. Front. Gyrus
R. Cerebellum
R. Sup. Temp. Gyrus
L. Inf. Temp. Gyrus
R. Mid. Front. Gyrus
R. Mid. Temp. Gyrus
L. Lingual Gyrus
R. Sup. Front. Gyrus
L. Mid. Temp. Gyrus
R. Lat. Forb. Gyrus
L. Fusiform Gyrus
L. Gyrus Rectus
L. Amygdala
R. Insular
R. Amygdala
R. Entorhinal Area
L. Sup. Front. Gyrus
R. Sup. Parietal Lobule
R. Angular Gyrus
R. Gyrus Rectus
L. Mid. Occipital Gyrus
R. Precuneus
L. Precuneus
L. Inf. Occipital Gyrus
L. Sup. Parietal Lobule
R. Red Nucleus
R. Inf. Cerebellar Peduncle
L. Medulla
L. Cerebellum WM
L. Fornix (Column&Body)
L. Tapatum
R. Tapatum
L. Lingual WM
R. Post. Thalamic Radiation
R. Substancia Nigra
L. Substancia Nigra
L. Post. Limb Int. Capsule
L. Sup. Occipital Gyrus
L. Rectus WM
R. Corticospinal Tract
R. Pontine Crossing Tract
R. Mid. Cerebellar Peduncle
L. Cerebral Peduncle
R. Mid. Forb. WM
R. Angular WM
L. Ant. Corona Radiata
R. Sup. Long. Fasciculus
L. Sup. Front. WM
R. Sup. Parietal WM
L. Postcent. WM
R. Precentral WM
L. Inf. Front. WM
R. Precuneus WM
L. Cing. (Hippocampus)
R. Mid. Temp. WM
L. Entorhinal Area
L. Midbrain
L. Lat. Forb. WM
R. Inf. Occipital WM
R. Inf. Front. WM
L. Fusiform WM
L. Body CC
R. Inf. Focc. Fasciculus
L. Ant. Limb Int. Capsule
L. Inf. Occipital WM
R. Cuneus WM
R. Cing. (Cingulate Gyrus)
L. Precuneus WM
R. Rectus WM
L. Rl. Pt. Int. Capsule
L. Supramarginal WM
R. Sup. Occipital WM
R. Supramarginal WM
L. Sup. Cerebellar Peduncle
L. Sup. Corona Radiata
R. Post. Limb Int. Capsule
L. Cingulate Gyrus
L. Mid. Forb. WM
R. Cuneus
R. Mid. Occipital WM
L. Cing.
L. Thalamus
L. Hippocampus
L. Fornix(cres) ST
L. Mid. Occipital WM
L. Mid. Temp. WM
R. Putamen
L. Globus Pallidus
Rescan
Motion
Sex
Interval
Baseline
p-value
-log p
50
25
10
2
0
coefficient
2 Std Dev
1 Std Dev
0
-1 Std Dev
-2 Std Dev
removed or p>0.05
coefficient
DTI Scalar: AD
Segmentation: EVE Type-1

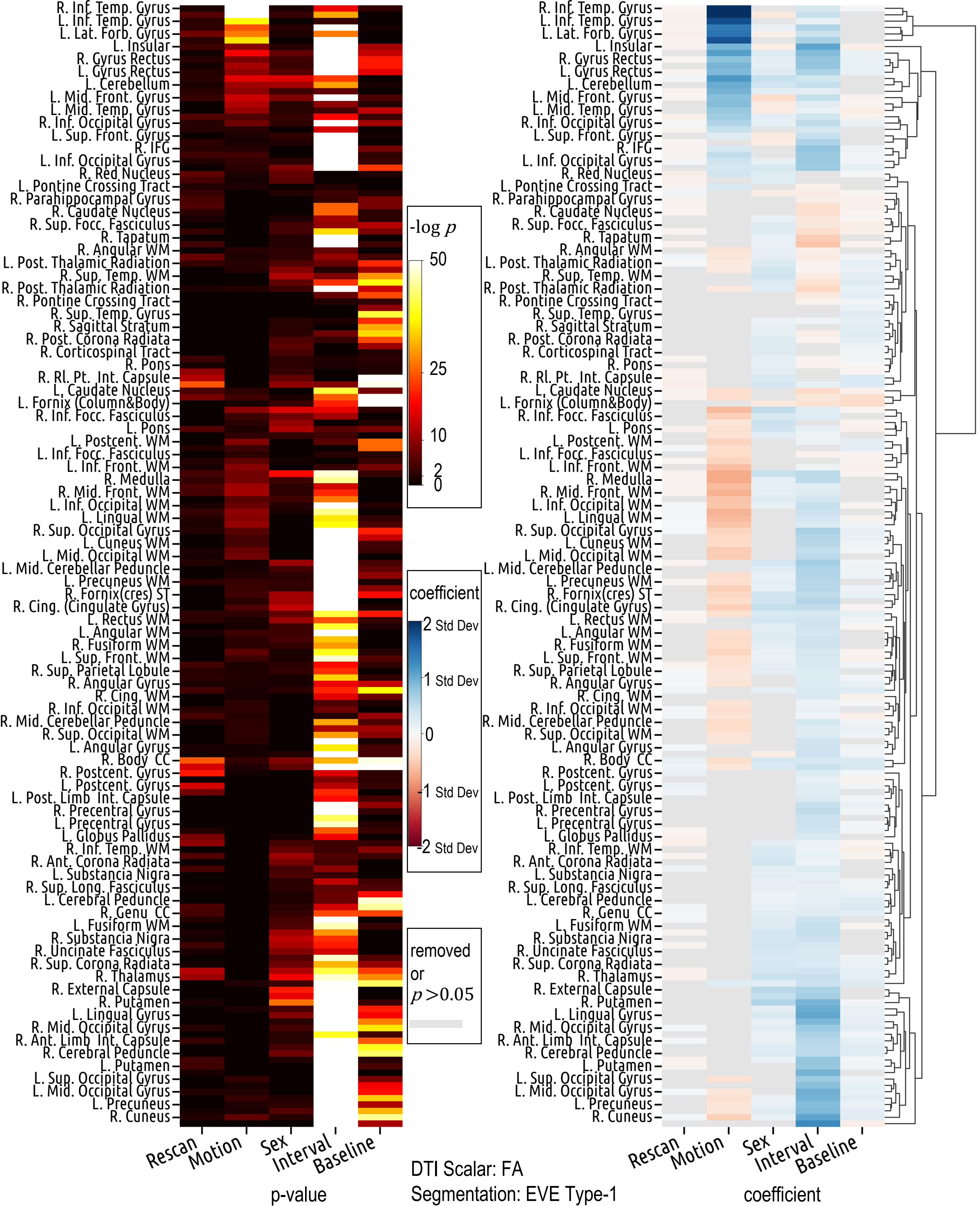
R. Inf. Temp. Gyrus
L. Inf. Temp. Gyrus
L. Lat. Forb. Gyrus
L. Insular
R. Gyrus Rectus
L. Gyrus Rectus
L. Cerebellum
L. Mid. Front. Gyrus
L. Mid. Temp. Gyrus
R. Inf. Occipital Gyrus
L. Sup. Front. Gyrus
R. IFG
L. Inf. Occipital Gyrus
R. Red Nucleus
L. Pontine Crossing Tract
R. Parahippocampal Gyrus
R. Caudate Nucleus
R. Sup. Focc. Fasciculus
R. Tapatum
R. Angular WM
L. Post. Thalamic Radiation
R. Sup. Temp. WM
R. Post. Thalamic Radiation
R. Pontine Crossing Tract
R. Sup. Temp. Gyrus
R. Sagittal Stratum
R. Post. Corona Radiata
R. Corticospinal Tract
R. Pons
R. Rl. Pt. Int. Capsule
L. Caudate Nucleus
L. Fornix (Column&Body)
R. Inf. Focc. Fasciculus
L. Pons
L. Postcent. WM
L. Inf. Focc. Fasciculus
L. Inf. Front. WM
R. Medulla
R. Mid. Front. WM
L. Inf. Occipital WM
L. Lingual WM
R. Sup. Occipital Gyrus
L. Cuneus WM
L. Mid. Occipital WM
L. Mid. Cerebellar Peduncle
L. Precuneus WM
R. Fornix(cres) ST
R. Cing. (Cingulate Gyrus)
L. Rectus WM
L. Angular WM
R. Fusiform WM
L. Sup. Front. WM
R. Sup. Parietal Lobule
R. Angular Gyrus
R. Cing. WM
R. Inf. Occipital WM
R. Mid. Cerebellar Peduncle
R. Sup. Occipital WM
L. Angular Gyrus
R. Body CC
R. Postcent. Gyrus
L. Postcent. Gyrus
L. Post. Limb Int. Capsule
R. Precentral Gyrus
L. Precentral Gyrus
L. Globus Pallidus
R. Inf. Temp. WM
R. Ant. Corona Radiata
L. Substancia Nigra
R. Sup. Long. Fasciculus
L. Cerebral Peduncle
R. Genu CC
L. Fusiform WM
R. Substancia Nigra
R. Uncinate Fasciculus
R. Sup. Corona Radiata
R. Thalamus
R. External Capsule
R. Putamen
L. Lingual Gyrus
R. Mid. Occipital Gyrus
R. Ant. Limb Int. Capsule
R. Cerebral Peduncle
L. Putamen
L. Sup. Occipital Gyrus
L. Mid. Occipital Gyrus
L. Precuneus
R. Cuneus
Rescan
Motion
Sex
Interval
Baseline
p-value
-log p
50
25
10
2
0
coefficient
2 Std Dev
1 Std Dev
0
-1 Std Dev
-2 Std Dev
removed
or
p>0.05
DTI Scalar: FA
Segmentation: EVE Type-1
coefficient

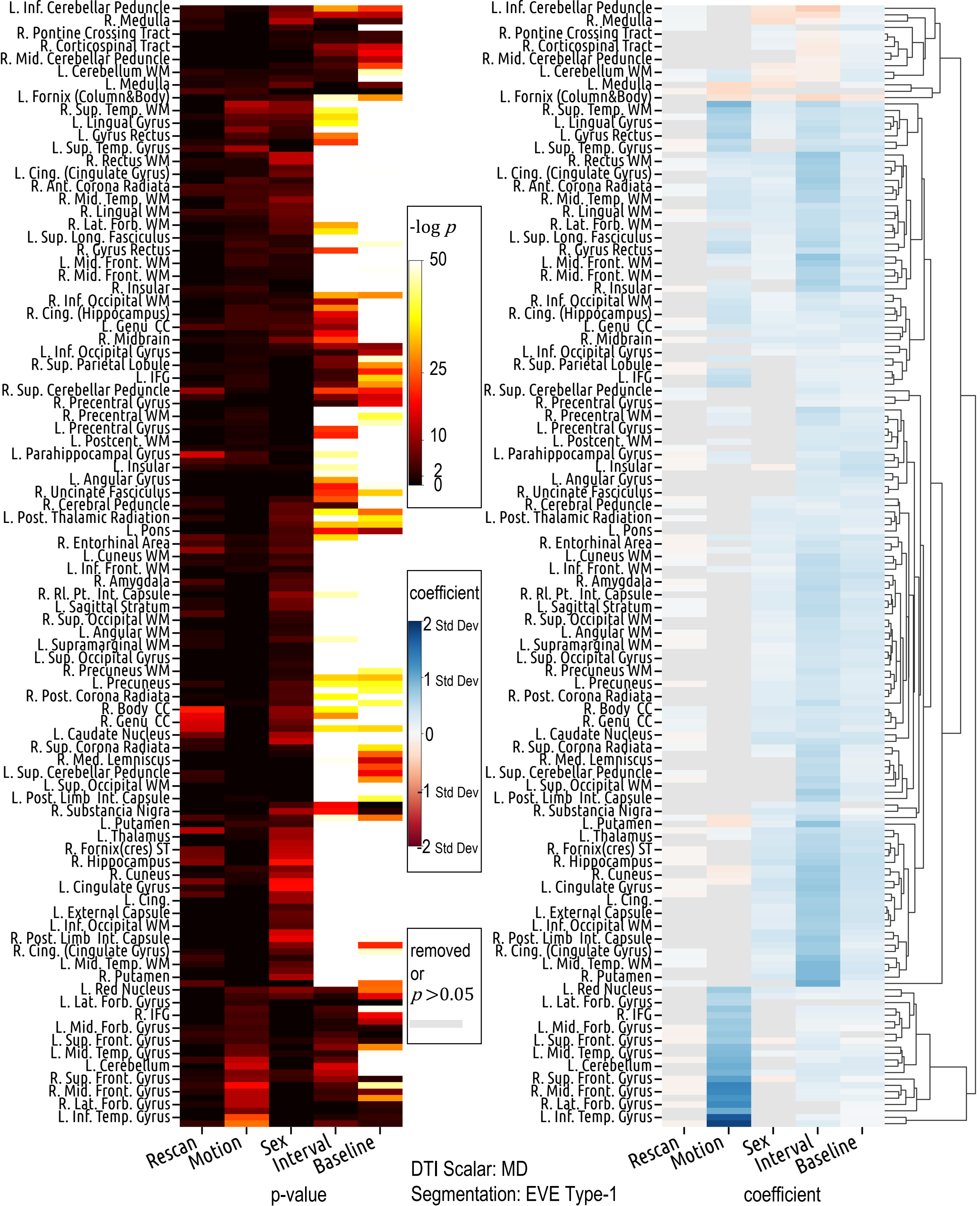
L. Inf. Cerebellar Peduncle
R. Medulla
R. Pontine Crossing Tract
R. Corticospinal Tract
R. Mid. Cerebellar Peduncle
L. Cerebellum WM
L. Medulla
L. Fornix (Column&Body)
R. Sup. Temp. WM
L. Lingual Gyrus
L. Gyrus Rectus
L. Sup. Temp. Gyrus
R. Rectus WM
L. Cing. (Cingulate Gyrus)
R. Ant. Corona Radiata
R. Mid. Temp. WM
R. Lingual WM
R. Lat. Forb. WM
L. Sup. Long. Fasciculus
R. Gyrus Rectus
L. Mid. Front. WM
R. Mid. Front. WM
R. Insular
R. Inf. Occipital WM
R. Cing. (Hippocampus)
L. Genu CC
R. Midbrain
L. Inf. Occipital Gyrus
R. Sup. Parietal Lobule
L. IFG
R. Sup. Cerebellar Peduncle
R. Precentral Gyrus
R. Precentral WM
L. Precentral Gyrus
L. Postcent. WM
L. Parahippocampal Gyrus
L. Insular
L. Angular Gyrus
R. Uncinate Fasciculus
R. Cerebral Peduncle
L. Post. Thalamic Radiation
L. Pons
R. Entorhinal Area
L. Cuneus WM
L. Inf. Front. WM
R. Amygdala
R. Rl. Pt. Int. Capsule
L. Sagittal Stratum
R. Sup. Occipital WM
L. Angular WM
L. Supramarginal WM
L. Sup. Occipital Gyrus
R. Precuneus WM
L. Precuneus
R. Post. Corona Radiata
R. Body CC
R. Genu CC
L. Caudate Nucleus
R. Sup. Corona Radiata
R. Med. Lemniscus
L. Sup. Cerebellar Peduncle
L. Sup. Occipital WM
L. Post. Limb Int. Capsule
R. Substancia Nigra
L. Putamen
L. Thalamus
R. Fornix(cres) ST
R. Hippocampus
R. Cuneus
L. Cingulate Gyrus
L. Cing.
L. External Capsule
L. Inf. Occipital WM
R. Post. Limb Int. Capsule
R. Cing. (Cingulate Gyrus)
L. Mid. Temp. WM
R. Putamen
L. Red Nucleus
L. Lat. Forb. Gyrus
R. IFG
L. Mid. Forb. Gyrus
L. Sup. Front. Gyrus
L. Mid. Temp. Gyrus
L. Cerebellum
R. Sup. Front. Gyrus
R. Mid. Front. Gyrus
R. Lat. Forb. Gyrus
L. Inf. Temp. Gyrus
Rescan
Motion
Sex
Interval
Baseline
p-value
-log p
50
25
10
2
0
coefficient
2 Std Dev
1 Std Dev
0
-1 Std Dev
-2 Std Dev
removed or p>0.05
DTI Scalar: MD
Segmentation: EVE Type-1
Rescan
Motion
Sex
Interval
Baseline
coefficient

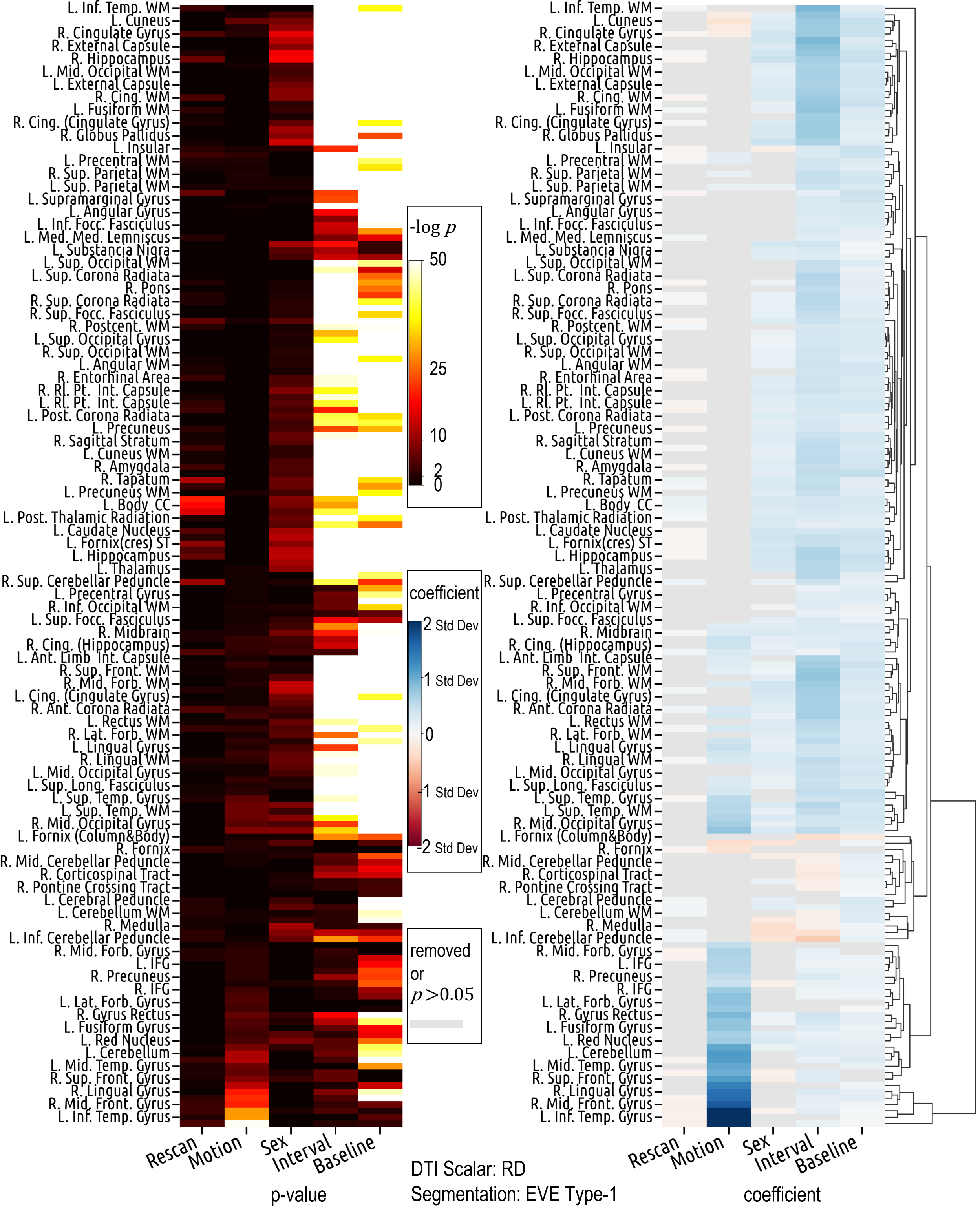

L. Inf. Temp. WM
L. Cuneus
R. Cingulate Gyrus
R. External Capsule
R. Hippocampus
L. Mid. Occipital WM
L. External Capsule
R. Cing. WM
L. Fusiform WM
R. Cing. (Cingulate Gyrus)
R. Globus Pallidus
L. Insular
L. Precentral WM
R. Sup. Parietal WM
L. Sup. Parietal WM
L. Supramarginal Gyrus
L. Angular Gyrus
L. Inf. Focc. Fasciculus
L. Med. Med. Lemniscus
L. Substancia Nigra
L. Sup. Occipital WM
L. Sup. Corona Radiata
R. Pons
R. Sup. Corona Radiata
R. Sup. Focc. Fasciculus
R. Postcent. WM
L. Sup. Occipital Gyrus
R. Sup. Occipital WM
L. Angular WM
R. Entorhinal Area
R. Rl. Pt. Int. Capsule
L. Rl. Pt. Int. Capsule
L. Post. Corona Radiata
L. Precuneus
R. Sagittal Stratum
L. Cuneus WM
R. Amygdala
R. Tapatum
L. Precuneus WM
L. Body CC
L. Post. Thalamic Radiation
L. Caudate Nucleus
L. Fornix(cres) ST
L. Hippocampus
L. Thalamus
R. Sup. Cerebellar Peduncle
L. Precentral Gyrus
R. Inf. Occipital WM
L. Sup. Focc. Fasciculus
R. Midbrain
R. Cing. (Hippocampus)
L. Ant. Limb Int. Capsule
R. Sup. Front. WM
R. Mid. Forb. WM
L. Cing. (Cingulate Gyrus)
R. Ant. Corona Radiata
L. Rectus WM
R. Lat. Forb. WM
L. Lingual Gyrus
R. Lingual WM
L. Mid. Occipital Gyrus
L. Sup. Long. Fasciculus
L. Sup. Temp. Gyrus
L. Sup. Temp. WM
R. Mid. Occipital Gyrus
L. Fornix (Column&Body)
R. Fornix
R. Mid. Cerebellar Peduncle
R. Corticospinal Tract
R. Pontine Crossing Tract
L. Cerebral Peduncle
L. Cerebellum WM
R. Medulla
L. Inf. Cerebellar Peduncle
R. Mid. Forb. Gyrus
L. IFG
R. Precuneus
R. IFG
L. Lat. Forb. Gyrus
R. Gyrus Rectus
L. Fusiform Gyrus
L. Red Nucleus
L. Cerebellum
L. Mid. Temp. Gyrus
R. Sup. Front. Gyrus
R. Lingual Gyrus
R. Mid. Front. Gyrus
L. Inf. Temp. Gyrus
Rescan
Motion
Sex
Interval
Baseline
p-value
-log p
50
25
10
2
0
coefficient
2 Std Dev
1 Std Dev
0
-1 Std Dev
-2 Std Dev
removed or p>0.05
coefficient
DTI Scalar: RD
Segmentation: EVE Type-1

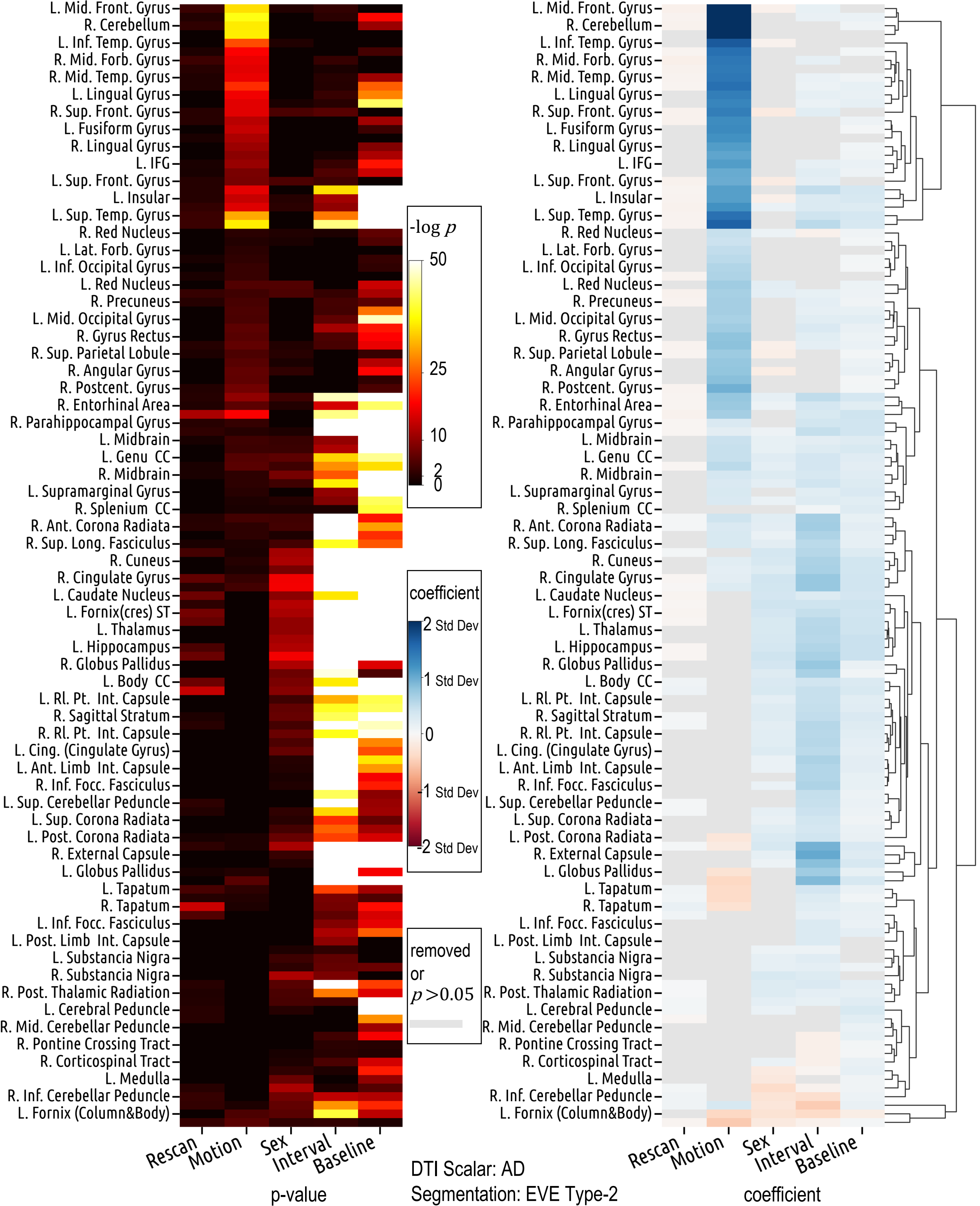

-log p
50
25
10
2
0
coefficient
2 Std Dev
1 Std Dev
0
-1 Std Dev
-2 Std Dev
removed or p>0.05
Rescan
Motion
Sex
Interval
Baseline
p-value
coefficient
DTI Scalar: AD
Segmentation: EVE Type-2
L. Mid. Front. Gyrus
R. Cerebellum
L. Inf. Temp. Gyrus
R. Mid. Forb. Gyrus
R. Mid. Temp. Gyrus
L. Lingual Gyrus
R. Sup. Front. Gyrus
L. Fusiform Gyrus
R. Lingual Gyrus
L. IFG
L. Sup. Front. Gyrus
L. Insular
L. Sup. Temp. Gyrus
R. Red Nucleus
L. Lat. Forb. Gyrus
L. Inf. Occipital Gyrus
L. Red Nucleus
R. Precuneus
L. Mid. Occipital Gyrus
R. Gyrus Rectus
R. Sup. Parietal Lobule
R. Angular Gyrus
R. Postcent. Gyrus
R. Entorhinal Area
R. Parahippocampal Gyrus
L. Midbrain
L. Genu CC
R. Midbrain
L. Supramarginal Gyrus
R. Splenium CC
R. Ant. Corona Radiata
R. Sup. Long. Fasciculus
R. Cuneus
R. Cingulate Gyrus
L. Caudate Nucleus
L. Fornix(cres) ST
L. Thalamus
L. Hippocampus
R. Globus Pallidus
L. Body CC
L. Rl. Pt. Int. Capsule
R. Sagittal Stratum
R. Rl. Pt. Int. Capsule
L. Cing. (Cingulate Gyrus)
L. Ant. Limb Int. Capsule
R. Inf. Focc. Fasciculus
L. Sup. Cerebellar Peduncle
L. Sup. Corona Radiata
L. Post. Corona Radiata
R. External Capsule
L. Globus Pallidus
L. Tapatum
R. Tapatum
L. Inf. Focc. Fasciculus
L. Post. Limb Int. Capsule
L. Substancia Nigra
R. Substancia Nigra
R. Post. Thalamic Radiation
L. Cerebral Peduncle
R. Mid. Cerebellar Peduncle
R. Pontine Crossing Tract
R. Corticospinal Tract
L. Medulla
R. Inf. Cerebellar Peduncle
L. Fornix (Column&Body)

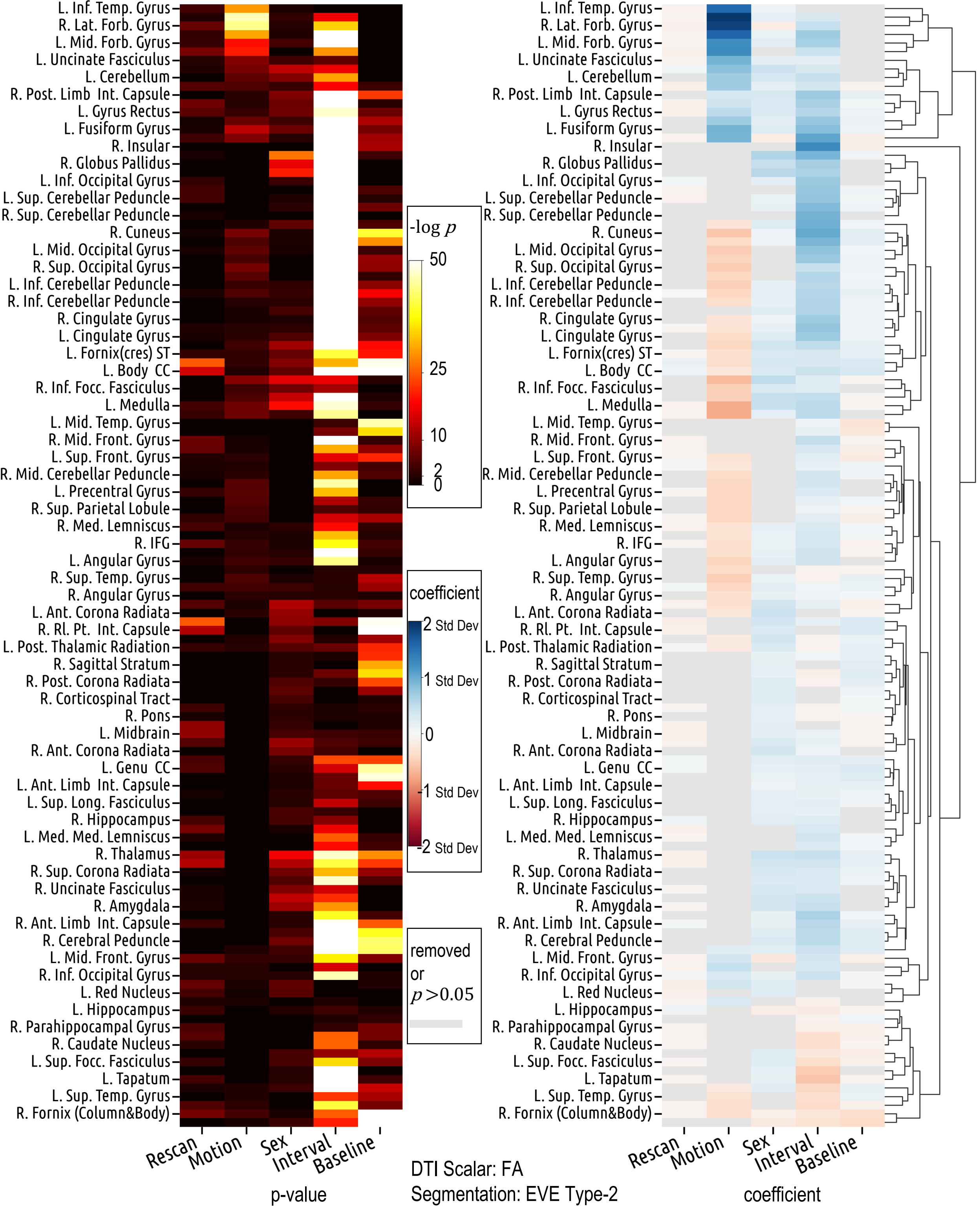

L. Inf. Temp. Gyrus
R. Lat. Forb. Gyrus
L. Mid. Forb. Gyrus
L. Uncinate Fasciculus
L. Cerebellum
R. Post. Limb Int. Capsule
L. Gyrus Rectus
L. Fusiform Gyrus
R. Insular
R. Globus Pallidus
L. Inf. Occipital Gyrus
L. Sup. Cerebellar Peduncle
R. Sup. Cerebellar Peduncle
R. Cuneus
L. Mid. Occipital Gyrus
R. Sup. Occipital Gyrus
L. Inf. Cerebellar Peduncle
R. Inf. Cerebellar Peduncle
R. Cingulate Gyrus
L. Cingulate Gyrus
L. Fornix(cres) ST
L. Body CC
R. Inf. Focc. Fasciculus
L. Medulla
L. Mid. Temp. Gyrus
R. Mid. Front. Gyrus
L. Sup. Front. Gyrus
R. Mid. Cerebellar Peduncle
L. Precentral Gyrus
R. Sup. Parietal Lobule
R. Med. Lemniscus
R. IFG
L. Angular Gyrus
R. Sup. Temp. Gyrus
R. Angular Gyrus
L. Ant. Corona Radiata
R. Rl. Pt. Int. Capsule
L. Post. Thalamic Radiation
R. Sagittal Stratum
R. Post. Corona Radiata
R. Corticospinal Tract
R. Pons
L. Midbrain
R. Ant. Corona Radiata
L. Genu CC
L. Ant. Limb Int. Capsule
L. Sup. Long. Fasciculus
R. Hippocampus
L. Med. Med. Lemniscus
R. Thalamus
R. Sup. Corona Radiata
R. Uncinate Fasciculus
R. Amygdala
R. Ant. Limb Int. Capsule
R. Cerebral Peduncle
L. Mid. Front. Gyrus
R. Inf. Occipital Gyrus
L. Red Nucleus
L. Hippocampus
R. Parahippocampal Gyrus
R. Caudate Nucleus
L. Sup. Focc. Fasciculus
L. Tapatum
L. Sup. Temp. Gyrus
R. Fornix (Column&Body)
Rescan
Motion
Sex
Interval
Baseline
p-value
-log p
50
25
10
2
0
coefficient
2 Std Dev
1 Std Dev
0
-1 Std Dev
-2 Std Dev
removed or p>0.05
DTI Scalar: FA
Segmentation: EVE Type-2
coefficient

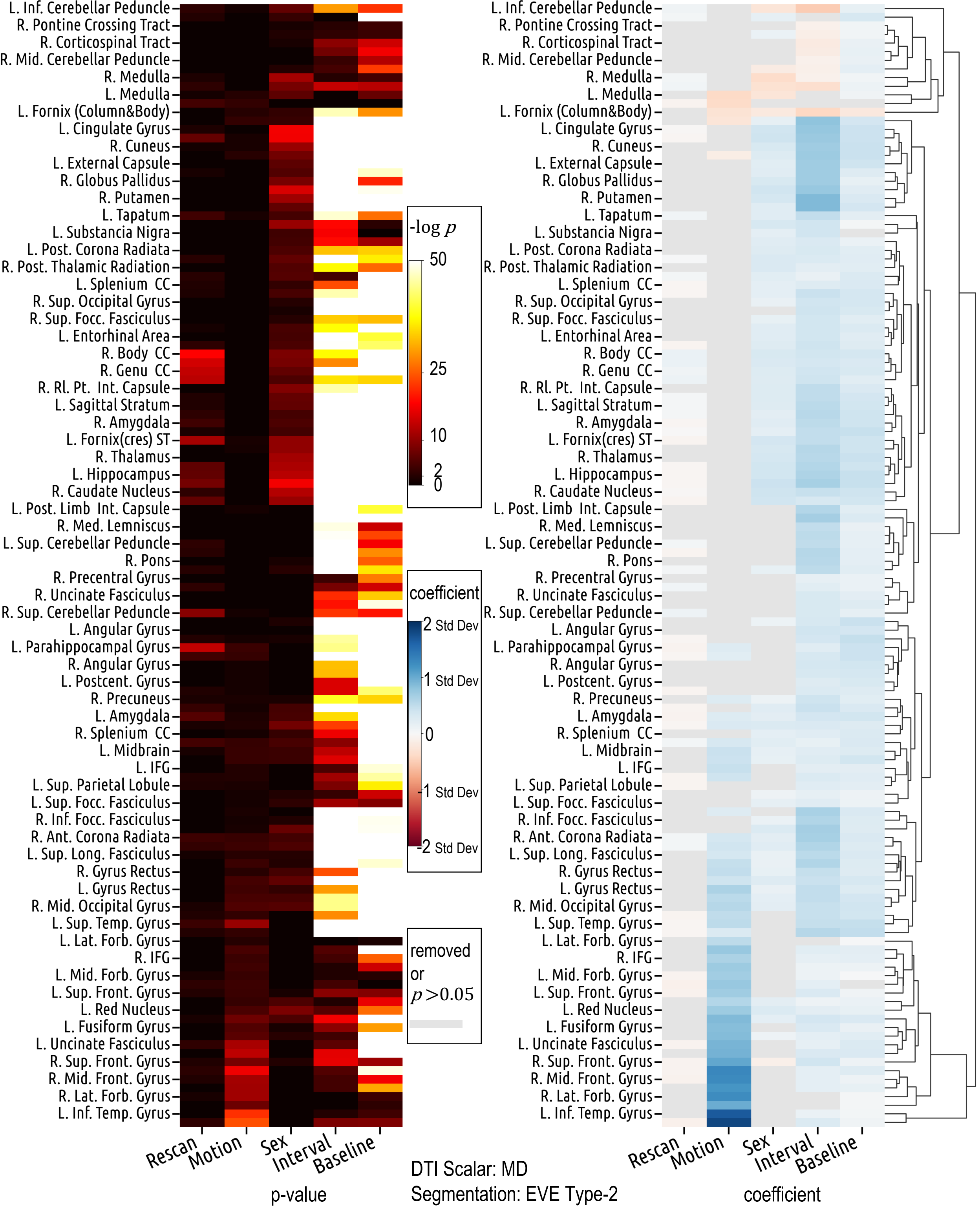
L. Inf. Cerebellar Peduncle
R. Pontine Crossing Tract
R. Corticospinal Tract
R. Mid. Cerebellar Peduncle
R. Medulla
L. Medulla
L. Fornix (Column&Body)
L. Cingulate Gyrus
R. Cuneus
L. External Capsule
R. Globus Pallidus
R. Putamen
L. Tapatum
L. Substancia Nigra
L. Post. Corona Radiata
R. Post. Thalamic Radiation
L. Splenium CC
R. Sup. Occipital Gyrus
R. Sup. Focc. Fasciculus
L. Entorhinal Area
R. Body CC
R. Genu CC
R. Rl. Pt. Int. Capsule
L. Sagittal Stratum
R. Amygdala
L. Fornix(cres) ST
R. Thalamus
L. Hippocampus
R. Caudate Nucleus
L. Post. Limb Int. Capsule
R. Med. Lemniscus
L. Sup. Cerebellar Peduncle
R. Pons
R. Precentral Gyrus
R. Uncinate Fasciculus
R. Sup. Cerebellar Peduncle
L. Angular Gyrus
L. Parahippocampal Gyrus
R. Angular Gyrus
L. Postcent. Gyrus
R. Precuneus
L. Amygdala
R. Splenium CC
L. Midbrain
L. IFG
L. Sup. Parietal Lobule
L. Sup. Focc. Fasciculus
R. Inf. Focc. Fasciculus
R. Ant. Corona Radiata
L. Sup. Long. Fasciculus
R. Gyrus Rectus
L. Gyrus Rectus
R. Mid. Occipital Gyrus
L. Sup. Temp. Gyrus
L. Lat. Forb. Gyrus
R. IFG
L. Mid. Forb. Gyrus
L. Sup. Front. Gyrus
L. Red Nucleus
L. Fusiform Gyrus
L. Uncinate Fasciculus
R. Sup. Front. Gyrus
R. Mid. Front. Gyrus
R. Lat. Forb. Gyrus
L. Inf. Temp. Gyrus
Rescan
Motion
Sex
Interval
Baseline
p-value
-log p
50
25
10
2
0
coefficient
2 Std Dev
1 Std Dev
0
-1 Std Dev
-2 Std Dev
removed or p>0.05
coefficient
DTI Scalar: MD
Segmentation: EVE Type-2

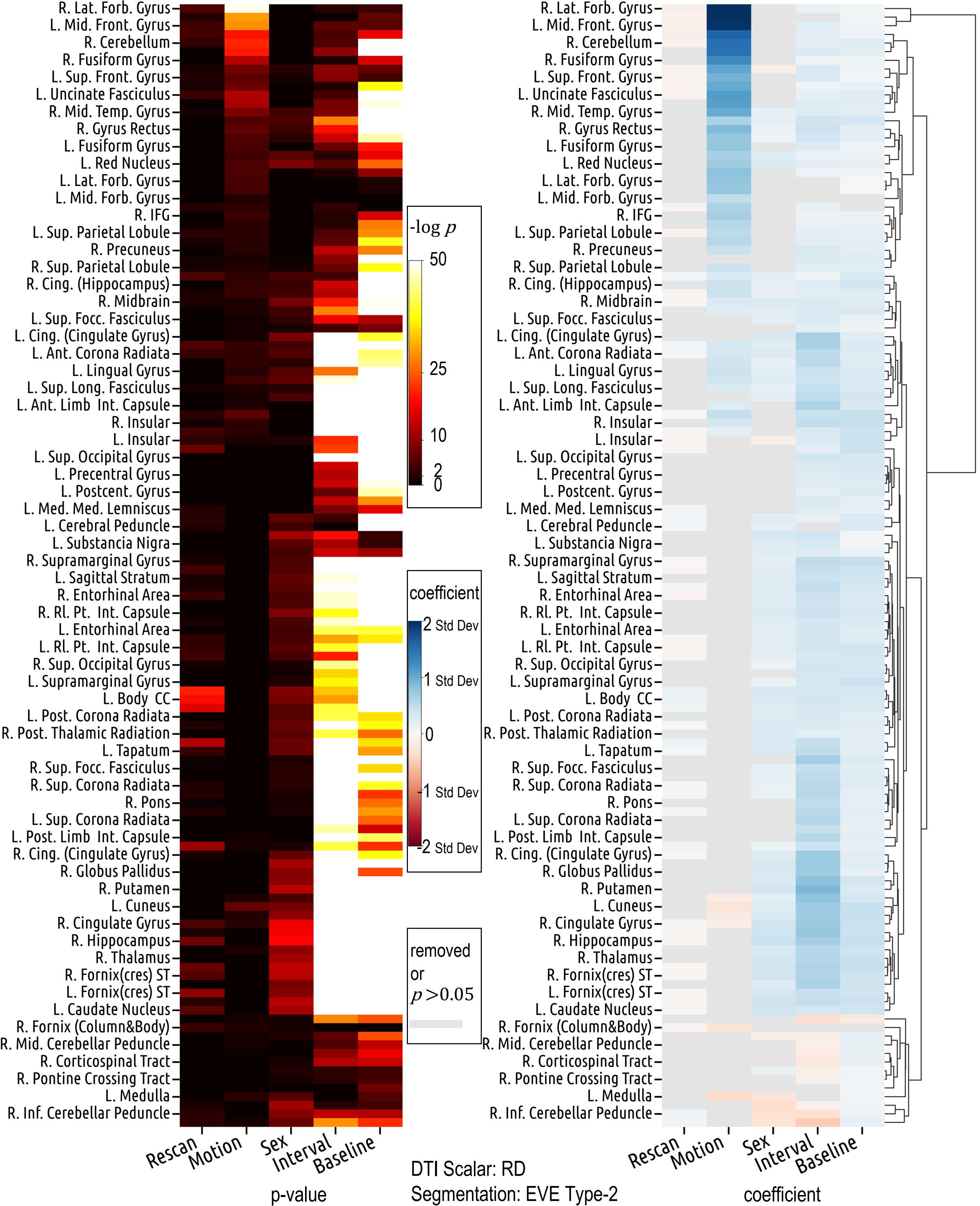

R. Lat. Forb. Gyrus
L. Mid. Front. Gyrus
R. Cerebellum
R. Fusiform Gyrus
L. Sup. Front. Gyrus
L. Uncinate Fasciculus
R. Mid. Temp. Gyrus
R. Gyrus Rectus
L. Fusiform Gyrus
L. Red Nucleus
L. Lat. Forb. Gyrus
L. Mid. Forb. Gyrus
R. IFG
L. Sup. Parietal Lobule
R. Precuneus
R. Sup. Parietal Lobule
R. Cing. (Hippocampus)
R. Midbrain
L. Sup. Focc. Fasciculus
L. Cing. (Cingulate Gyrus)
L. Ant. Corona Radiata
L. Lingual Gyrus
L. Sup. Long. Fasciculus
L. Ant. Limb Int. Capsule
R. Insular
L. Insular
L. Sup. Occipital Gyrus
L. Precentral Gyrus
L. Postcent. Gyrus
L. Med. Med. Lemniscus
L. Cerebral Peduncle
L. Substancia Nigra
R. Supramarginal Gyrus
L. Sagittal Stratum
R. Entorhinal Area
R. Rl. Pt. Int. Capsule
L. Entorhinal Area
L. Rl. Pt. Int. Capsule
R. Sup. Occipital Gyrus
L. Supramarginal Gyrus
L. Body CC
L. Post. Corona Radiata
R. Post. Thalamic Radiation
L. Tapatum
R. Sup. Focc. Fasciculus
R. Sup. Corona Radiata
R. Pons
L. Sup. Corona Radiata
L. Post. Limb Int. Capsule
R. Cing. (Cingulate Gyrus)
R. Globus Pallidus
R. Putamen
L. Cuneus
R. Cingulate Gyrus
R. Hippocampus
R. Thalamus
R. Fornix(cres) ST
L. Fornix(cres) ST
L. Caudate Nucleus
R. Fornix (Column&Body)
R. Mid. Cerebellar Peduncle
R. Corticospinal Tract
R. Pontine Crossing Tract
L. Medulla
R. Inf. Cerebellar Peduncle
Rescan
Motion
Sex
Interval
Baseline
p-value
-log p
50
25
10
2
0
coefficient
2 Std Dev
1 Std Dev
0
-1 Std Dev
-2 Std Dev
removed or p>0.05
DTI Scalar: RD
Segmentation: EVE Type-2
coefficient

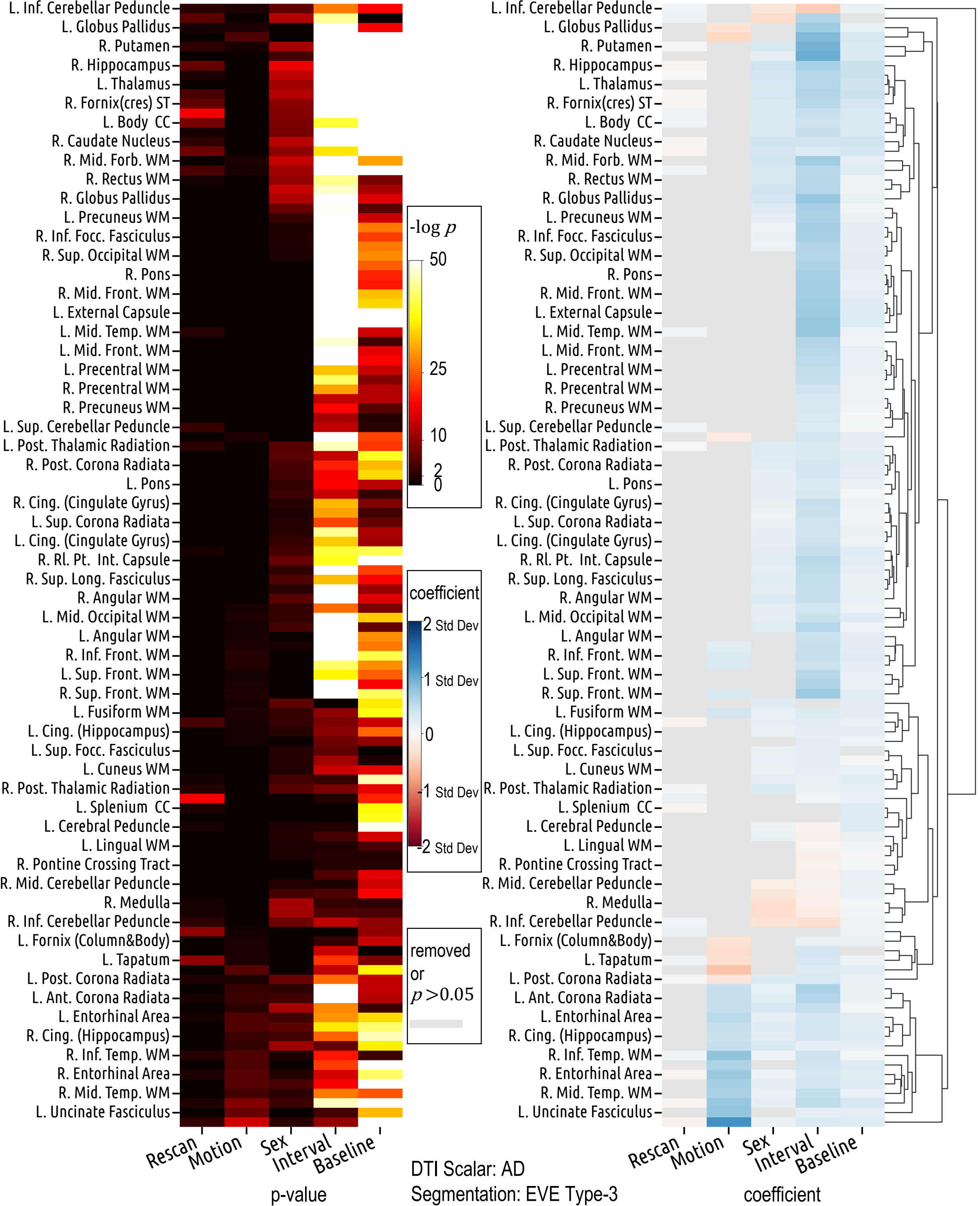

L. Inf. Cerebellar Peduncle
L. Globus Pallidus
R. Putamen
R. Hippocampus
L. Thalamus
R. Fornix(cres) ST
L. Body CC
R. Caudate Nucleus
R. Mid. Forb. WM
R. Rectus WM
R. Globus Pallidus
L. Precuneus WM
R. Inf. Focc. Fasciculus
R. Sup. Occipital WM
R. Pons
R. Mid. Front. WM
L. External Capsule
L. Mid. Temp. WM
L. Mid. Front. WM
L. Precentral WM
R. Precentral WM
R. Precuneus WM
L. Sup. Cerebellar Peduncle
L. Post. Thalamic Radiation
R. Post. Corona Radiata
L. Pons
R. Cing. (Cingulate Gyrus)
L. Sup. Corona Radiata
L. Cing. (Cingulate Gyrus)
R. Rl. Pt. Int. Capsule
R. Sup. Long. Fasciculus
R. Angular WM
L. Mid. Occipital WM
L. Angular WM
R. Inf. Front. WM
L. Sup. Front. WM
R. Sup. Front. WM
L. Fusiform WM
L. Cing. (Hippocampus)
L. Sup. Focc. Fasciculus
L. Cuneus WM
R. Post. Thalamic Radiation
L. Splenium CC
L. Cerebral Peduncle
L. Lingual WM
R. Pontine Crossing Tract
R. Mid. Cerebellar Peduncle
R. Medulla
R. Inf. Cerebellar Peduncle
L. Fornix (Column&Body)
L. Tapatum
L. Post. Corona Radiata
L. Ant. Corona Radiata
L. Entorhinal Area
R. Cing. (Hippocampus)
R. Inf. Temp. WM
R. Entorhinal Area
R. Mid. Temp. WM
L. Uncinate Fasciculus
Rescan
Motion
Sex
Interval
Baseline
p-value
-log p
50
25
10
2
0
coefficient
2 Std Dev
1 Std Dev
0
-1 Std Dev
-2 Std Dev
removed or p>0.05
DTI Scalar: AD
Segmentation: EVE Type-3
coefficient

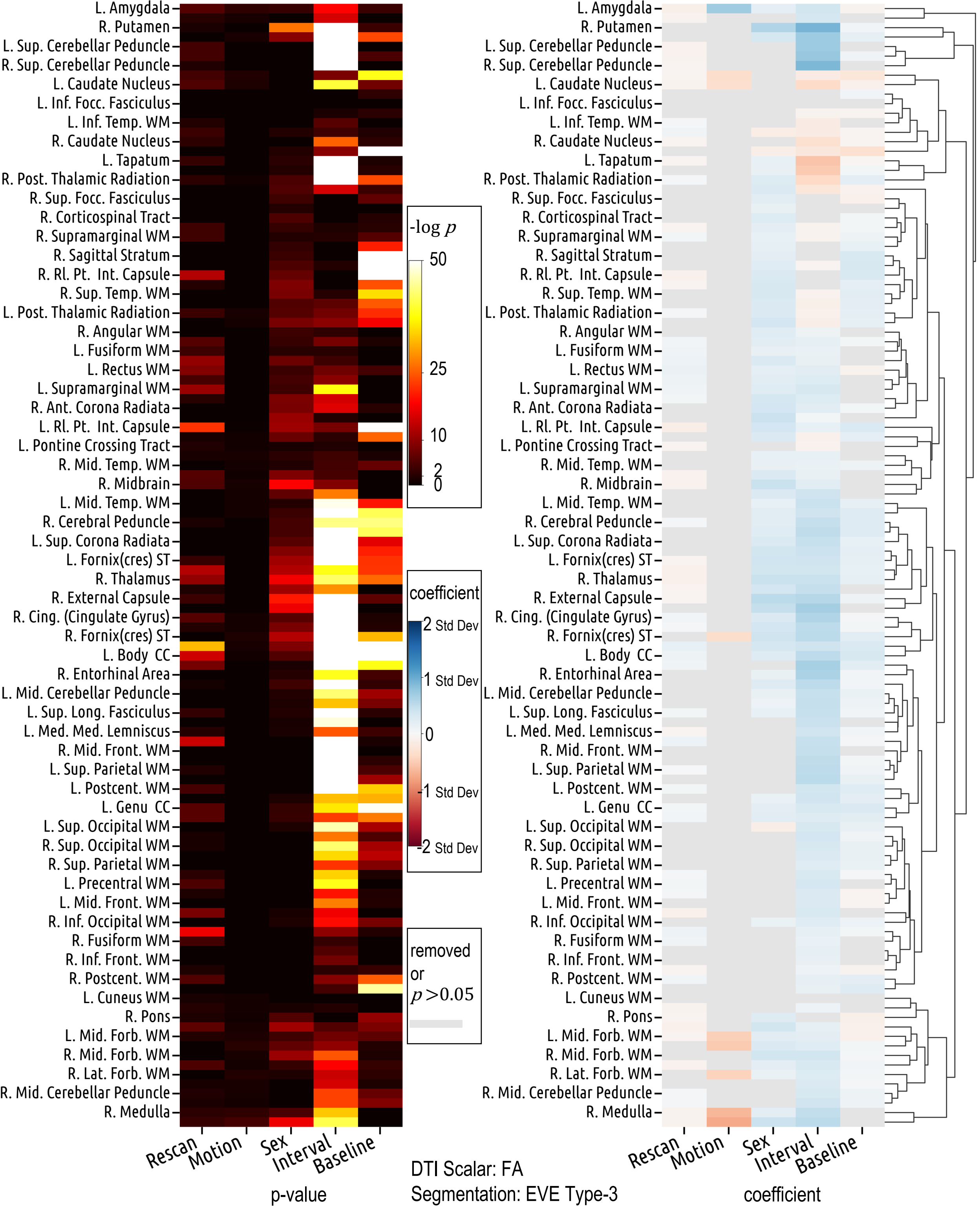

L. Amygdala
R. Putamen
L. Sup. Cerebellar Peduncle
R. Sup. Cerebellar Peduncle
L. Caudate Nucleus
L. Inf. Focc. Fasciculus
L. Inf. Temp. WM
R. Caudate Nucleus
L. Tapatum
R. Post. Thalamic Radiation
R. Sup. Focc. Fasciculus
R. Corticospinal Tract
R. Supramarginal WM
R. Sagittal Stratum
R. Rl. Pt. Int. Capsule
R. Sup. Temp. WM
L. Post. Thalamic Radiation
R. Angular WM
L. Fusiform WM
L. Rectus WM
L. Supramarginal WM
R. Ant. Corona Radiata
L. Rl. Pt. Int. Capsule
L. Pontine Crossing Tract
R. Mid. Temp. WM
R. Midbrain
L. Mid. Temp. WM
R. Cerebral Peduncle
L. Sup. Corona Radiata
L. Fornix(cres) ST
R. Thalamus
R. External Capsule
R. Cing. (Cingulate Gyrus)
R. Fornix(cres) ST
L. Body CC
R. Entorhinal Area
L. Mid. Cerebellar Peduncle
L. Sup. Long. Fasciculus
L. Med. Med. Lemniscus
R. Mid. Front. WM
L. Sup. Parietal WM
L. Postcent. WM
L. Genu CC
L. Sup. Occipital WM
R. Sup. Occipital WM
R. Sup. Parietal WM
L. Precentral WM
L. Mid. Front. WM
R. Inf. Occipital WM
R. Fusiform WM
R. Inf. Front. WM
R. Postcent. WM
L. Cuneus WM
R. Pons
L. Mid. Forb. WM
R. Mid. Forb. WM
R. Lat. Forb. WM
R. Mid. Cerebellar Peduncle
R. Medulla
Rescan
Motion
Sex
Interval
Baseline
p-value
-log p
50
25
10
2
0
coefficient
2 Std Dev
1 Std Dev
0
-1 Std Dev
-2 Std Dev
removed or p>0.05
DTI Scalar: FA
Segmentation: EVE Type-3
coefficient

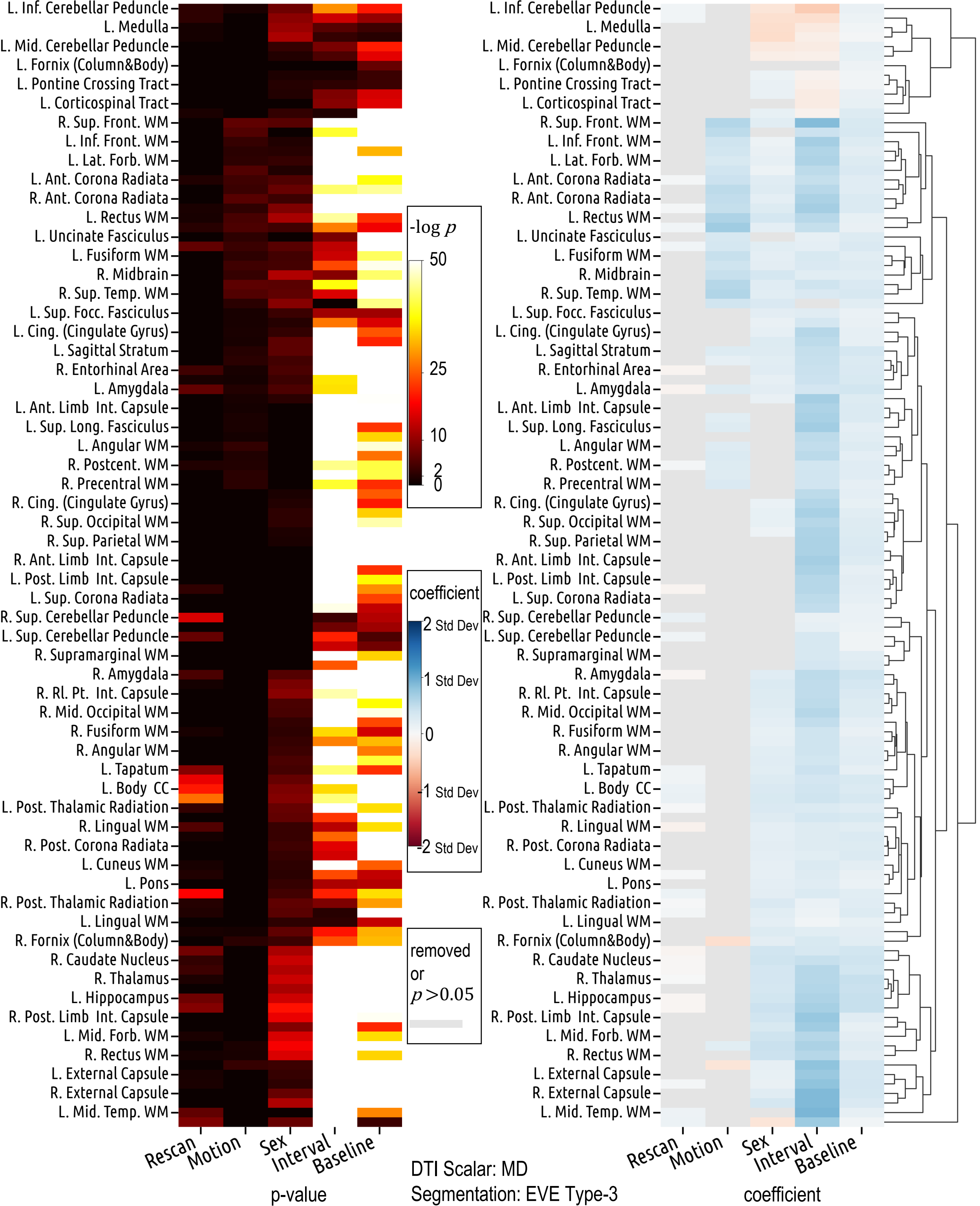

L. Inf. Cerebellar Peduncle
L. Medulla
L. Mid. Cerebellar Peduncle
L. Fornix (Column&Body)
L. Pontine Crossing Tract
L. Corticospinal Tract
R. Sup. Front. WM
L. Inf. Front. WM
L. Lat. Forb. WM
L. Ant. Corona Radiata
R. Ant. Corona Radiata
L. Rectus WM
L. Uncinate Fasciculus
L. Fusiform WM
R. Midbrain
R. Sup. Temp. WM
L. Sup. Focc. Fasciculus
L. Cing. (Cingulate Gyrus)
L. Sagittal Stratum
R. Entorhinal Area
L. Amygdala
L. Ant. Limb Int. Capsule
L. Sup. Long. Fasciculus
L. Angular WM
R. Postcent. WM
R. Precentral WM
R. Cing. (Cingulate Gyrus)
R. Sup. Occipital WM
R. Sup. Parietal WM
R. Ant. Limb Int. Capsule
L. Post. Limb Int. Capsule
L. Sup. Corona Radiata
R. Sup. Cerebellar Peduncle
L. Sup. Cerebellar Peduncle
R. Supramarginal WM
R. Amygdala
R. Rl. Pt. Int. Capsule
R. Mid. Occipital WM
R. Fusiform WM
R. Angular WM
L. Tapatum
L. Body CC
L. Post. Thalamic Radiation
R. Lingual WM
R. Post. Corona Radiata
L. Cuneus WM
L. Pons
R. Post. Thalamic Radiation
L. Lingual WM
R. Fornix (Column&Body)
R. Caudate Nucleus
R. Thalamus
L. Hippocampus
R. Post. Limb Int. Capsule
L. Mid. Forb. WM
R. Rectus WM
L. External Capsule
R. External Capsule
L. Mid. Temp. WM
-log p
50
25
10
2
0
coefficient
2 Std Dev
1 Std Dev
0
-1 Std Dev
-2 Std Dev
removed or p>0.05
Rescan
Motion
Sex
Interval
Baseline
p-value
DTI Scalar: MD
Segmentation: EVE Type-3
coefficient

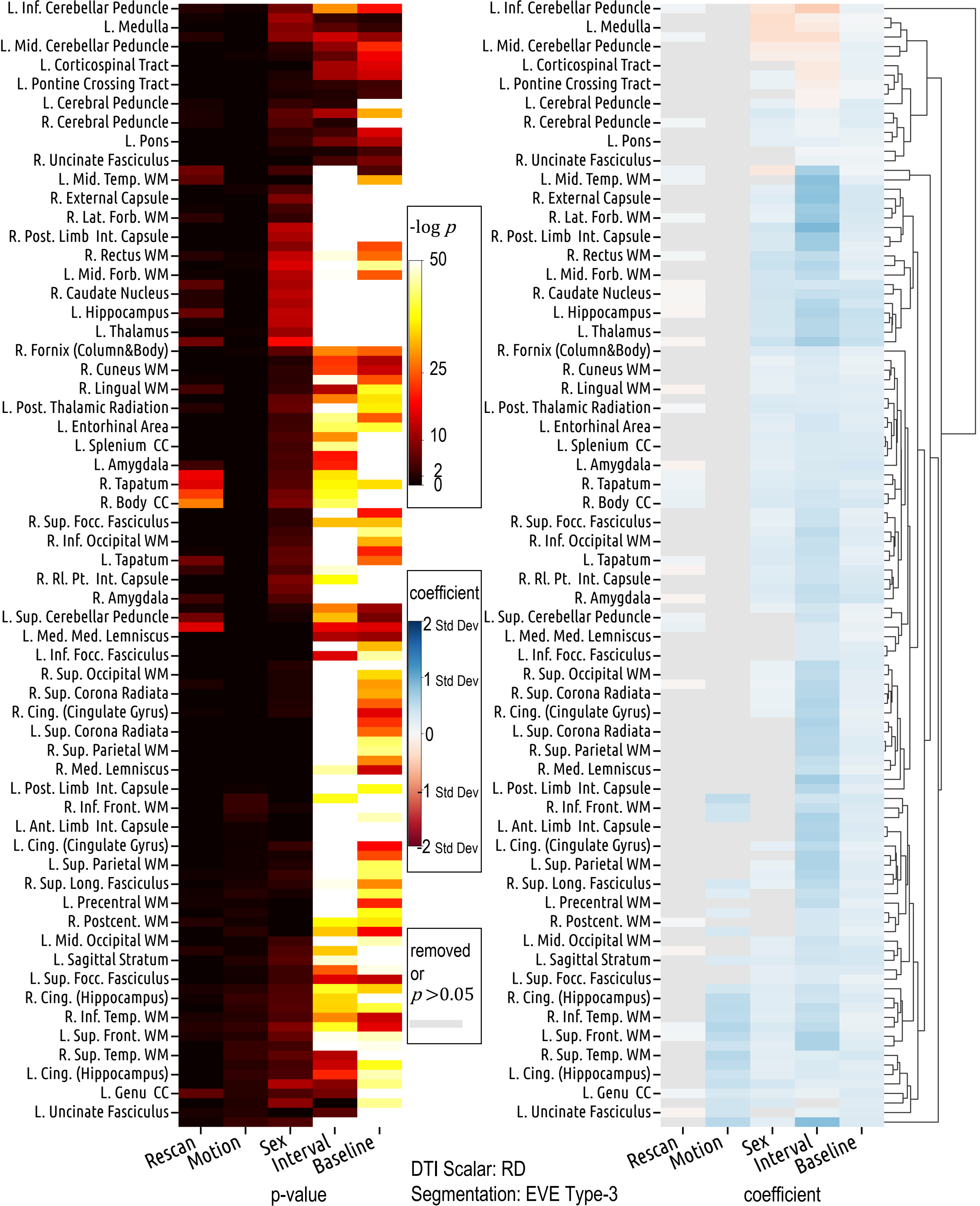
L. Inf. Cerebellar Peduncle
L. Medulla
L. Mid. Cerebellar Peduncle
L. Corticospinal Tract
L. Pontine Crossing Tract
L. Cerebral Peduncle
R. Cerebral Peduncle
L. Pons
R. Uncinate Fasciculus
L. Mid. Temp. WM
R. External Capsule
R. Lat. Forb. WM
R. Post. Limb Int. Capsule
R. Rectus WM
L. Mid. Forb. WM
R. Caudate Nucleus
L. Hippocampus
L. Thalamus
R. Fornix (Column&Body)
R. Cuneus WM
R. Lingual WM
L. Post. Thalamic Radiation
L. Entorhinal Area
L. Splenium CC
L. Amygdala
R. Tapatum
R. Body CC
R. Sup. Focc. Fasciculus
R. Inf. Occipital WM
L. Tapatum
R. Rl. Pt. Int. Capsule
R. Amygdala
L. Sup. Cerebellar Peduncle
L. Med. Med. Lemniscus
L. Inf. Focc. Fasciculus
R. Sup. Occipital WM
R. Sup. Corona Radiata
R. Cing. (Cingulate Gyrus)
L. Sup. Corona Radiata
R. Sup. Parietal WM
R. Med. Lemniscus
L. Post. Limb Int. Capsule
R. Inf. Front. WM
L. Ant. Limb Int. Capsule
L. Cing. (Cingulate Gyrus)
L. Sup. Parietal WM
R. Sup. Long. Fasciculus
L. Precentral WM
R. Postcent. WM
L. Mid. Occipital WM
L. Sagittal Stratum
L. Sup. Focc. Fasciculus
R. Cing. (Hippocampus)
R. Inf. Temp. WM
L. Sup. Front. WM
R. Sup. Temp. WM
L. Cing. (Hippocampus)
L. Genu CC
L. Uncinate Fasciculus
Rescan
Motion
Sex
Interval
Baseline
p-value
-log $p$
50
25
10
2
0
coefficient
2 Std Dev
1 Std Dev
0
-1 Std Dev
-2 Std Dev
removed or $p > 0.05$
DTI Scalar: RD
Segmentation: EVE Type-3
coefficient
