## Supplemental Figures - Part 2 for "Characterizing patterns of diffusion tensor imaging variance in aging brains"

**Supplementary Materials Part 2**

| **# Subjects by race** | **Female** | **Male** | **Combined** |
| --- | --- | --- | --- |
| White | 348 | 346 | 694 |
| Black or African American | 163 | 92 | 255 |
| American Indian or Alaska Native | 2 | 2 | 4 |
| Asian | 16 | 2 | 18 |
| Other (or unavailable) | 33 | 31 | 64 |
| Total | 562 | 473 | 1035 |

**Table S1** Number of subjects of each race category for the BLSA data we included.

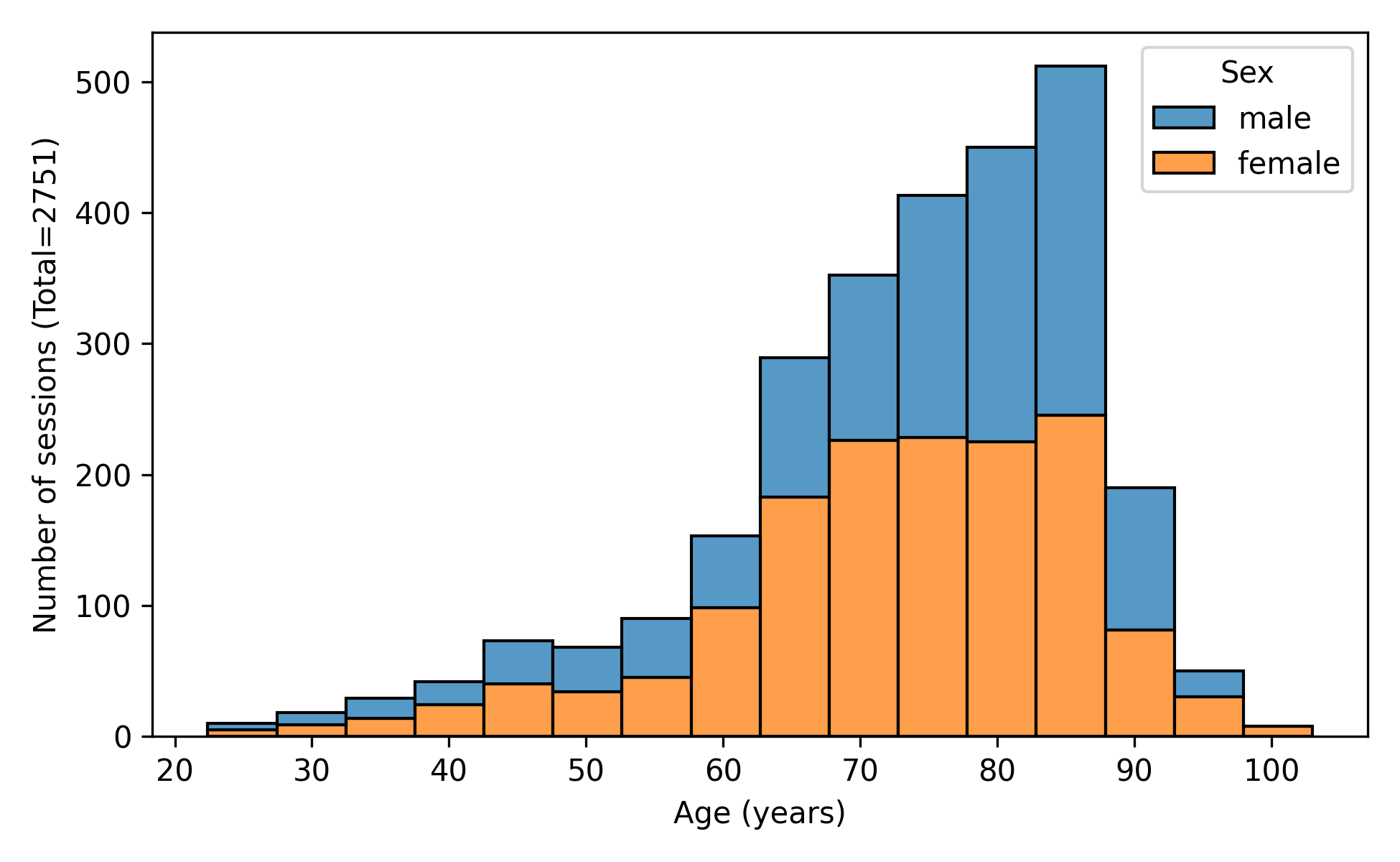
**Figure S1** Age distribution of the sessions of the BLSA subjects included for this study.

| **Dataset** | **# Subjects**  **(Females / Males)** | **# Sessions**  **(Females / Males)** | **Age range (years)** | **Age mean ± std (years)** |
| --- | --- | --- | --- | --- |
| BLSA | 1007 (551/456) | 2577 (1438/1139) | 22.4 - 101.0 | 72.7 ± 13.5 |
| ADNI | 652 (389/263) | 1370 (807/563) | 50.5 - 95.4 | 73.5 ± 7.2 |
| BIOCARD | 149 (96/53) | 336 (217/119) | 34.0 - 93.0 | 70.4 ± 7.9 |
| Total | 1808 (1036/772) | 4283 (2462/1821) | 22.4 - 101.0 | 72.8 ± 11.5 |

**Table S2** The datasets used for validating our findings.

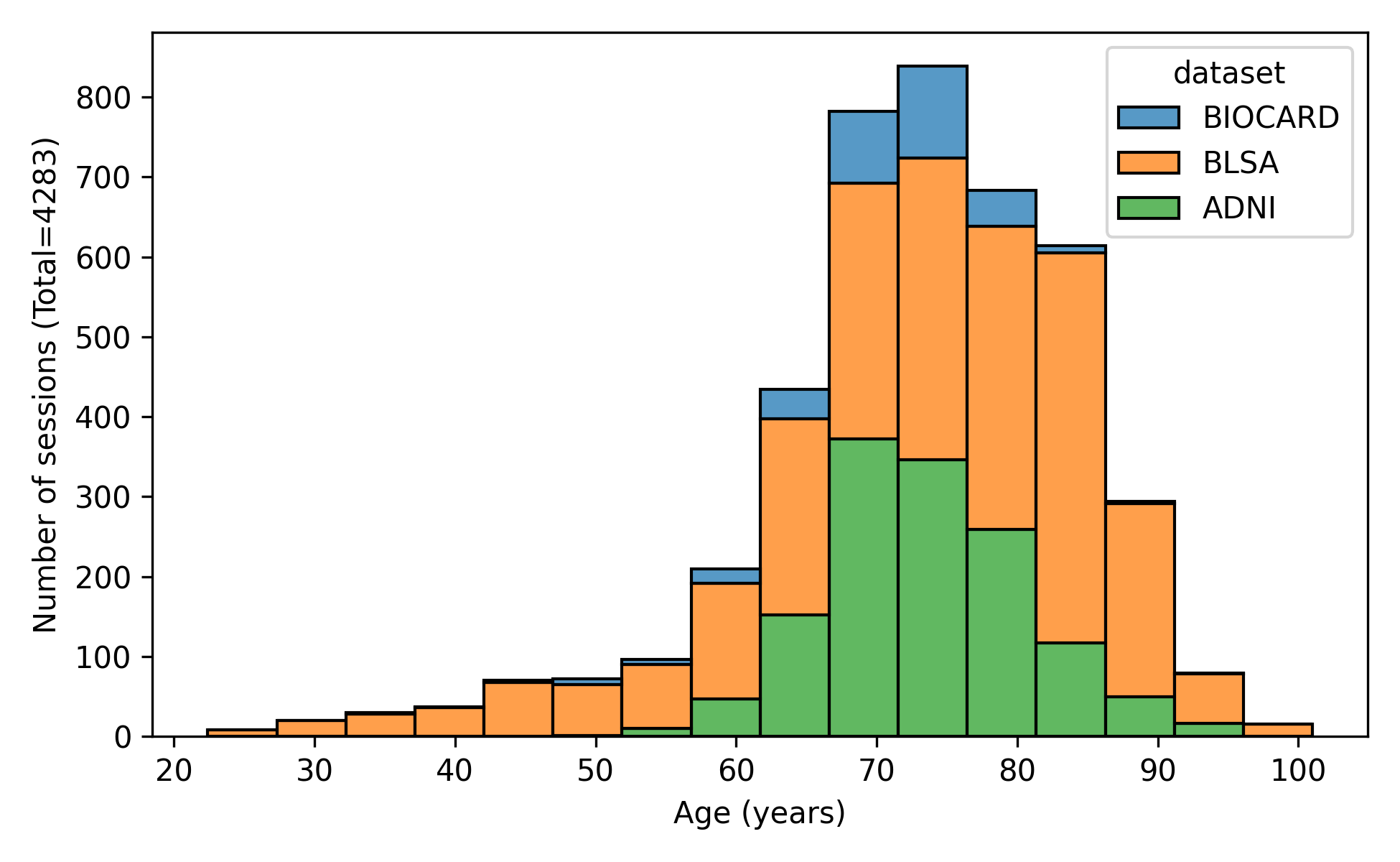

**Figure S2** Age distribution of the datasets used for validating our findings.
